# Cost-effectiveness of overseas testing and treatment for tuberculosis infection among United States-bound refugees: a mathematical modelling analysis

**DOI:** 10.64898/2026.01.27.26344963

**Authors:** Yuli Lily Hsieh, Christina R Phares, Suzanne M Marks, Brian Maskery, Garrett R Beeler Asay, Susan A Maloney, Nicole A Swartwood, Anand Date, Ted Cohen, Nicolas A Menzies

## Abstract

**Research in context:** *Evidence before this study:* Previous cost-effectiveness analyses have examined tuberculosis (TB) infection testing and latent TB infection (LTBI) treatment among migrants in high-income, low TB-incidence countries, including the United States, Canada, and Australia. These studies found that cost-effectiveness varied by setting, population risk, and intervention design. Refugees and asylum seekers—populations with higher TB exposure and reduced healthcare engagement post-arrival—were identified as high-priority groups. Studies suggested that diagnosis and treatment of LTBI before migrants depart their origin country could improve retention along the care cascade and yield better health and economic outcomes compared to post-arrival interventions. A recent pilot study demonstrated the feasibility of pre-departure TB infection testing and voluntary LTBI treatment among U.S.-bound immigrants in Vietnam. However, a cost-effectiveness analysis examining the addition of pre-departure LTBI treatment to pre-departure testing and post-arrival LTBI treatment for refugee and asylee populations is lacking.

*Added value of this study:* This model-based cost-effectiveness analysis extends prior work by evaluating the addition of pre-departure LTBI treatment to pre-departure testing and post-arrival LTBI treatment. It demonstrates that a pre-departure offer of LTBI treatment could increase overall treatment completion and enhance both health outcomes and cost-effectiveness.

*Implications of all the available evidence:* In many settings, recently arrived refugees have some of the highest risks of developing TB disease. Therefore, identifying preventive interventions that can reduce TB risk among the refugee population—in ways that are cost-effective, feasible, and respectful of individual autonomy—is a high public health priority. This study found that pre-departure TB infection testing and voluntary LTBI treatment would be a cost-effective addition to current post-arrival prevention approaches, reducing TB risk for a traditionally underserved population at high risk of TB disease.

**Background:** In the United States, preventing TB among refugee populations is a public health priority. We assessed the health impact and cost-effectiveness of strategies to diagnose and treat latent TB infection (LTBI) among U.S.-bound refugees from high TB incidence countries.

**Methods:** Using mathematical modelling, we simulated TB-related health outcomes and costs (2023 USD) among individuals entering the United States as refugees, from pre-departure medical evaluation until death. LTBI diagnosis was made via interferon-gamma release assay (IGRA), after ruling out TB disease. We compared three intervention strategies: (1) pre-departure IGRA testing for children (2-14 years) and post-arrival IGRA testing for adults (>14 years), with LTBI treatment offered in the United States; (2) pre-departure IGRA testing for children and adults, with LTBI treatment offered post-arrival; (3) pre-departure IGRA testing for children and adults with LTBI treatment offered pre-departure, then re-offered in the United States for individuals not completing treatment before U.S. arrival.

**Findings:** The intervention strategies were projected to avert 32-60% lifetime TB cases for children and adults, compared to no IGRA testing or LTBI treatment (‘no intervention’). Compared to Strategies 1 and 2, Strategy 3 produced greater health gains with lower incremental costs. Compared to no intervention, Strategy 3 had an incremental cost-effectiveness ratio of $45,000 per QALY gained for children, and $21,111 per QALY gained for adults.

**Interpretation:** Pre-departure IGRA testing and voluntary LTBI treatment could be cost-effective for preventing TB disease among U.S.-bound refugees, when provided in conjunction with existing services to diagnosis and treat TB disease.

**Funding:** CDC.

## INTRODUCTION

Global migration from high tuberculosis (TB) burden countries has played an important role in shaping TB epidemiology in low burden countries, including the United States.^1^ In 2023, 76% of TB cases in the United States occurred in non-U.S.-born individuals,^2^ among whom over 80% likely resulted from latent tuberculosis infection (LTBI) acquired before migration.^3^ Diagnosis and treatment of LTBI for non-U.S.-born individuals is therefore a key TB elimination strategy in the United States.^1,4,5^ Among international migrants, refugees face particularly high risks of TB due to multiple factors, including crowded living conditions and inadequate treatment access.^6,7^

Under the U.S. Immigration and Nationality Act, overseas medical examination for TB disease is mandatory for all U.S.-bound refugees, and individuals diagnosed with infectious TB disease must complete TB treatment before departure for the United States.^8^ Prior to 2024, as part of TB disease screening, children 2-14 years old examined in countries with WHO-estimated TB incidence ≥20 cases per 100,000 (hereinafter ‘high-TB-burden’ countries) were tested with an interferon gamma release assay (IGRA). The updated 2024 CDC Technical Instructions for Panel Physicians states that pre-departure IGRA testing is required for all applicants ≥2 years old examined in high-TB-burden countries.^9^ For applicants testing IGRA-positive and not diagnosed with TB disease, LTBI treatment is not required prior to U.S. entry though is recommended after arrival.^10^ While non-refugee immigrants pay for their own medical examinations, the U.S. government pays for these examinations and all required tests (e.g., IGRA) for refugees.

For refugees arriving in the United States over 2013-2023, 33% were children 2-14 years old. Among those diagnosed with LTBI and recommended to receive LTBI treatment, 65% initiated treatment, among whom 61% completed treatment (i.e., 40% of those recommended to receive treatment), based on CDC Electronic Disease Notification [EDN] data.^11^ One potential strategy to improve LTBI treatment uptake and completion is to offer treatment before U.S. entry. A recent pilot study demonstrated the feasibility of pre-departure IGRA testing and voluntary LTBI treatment among U.S.-bound immigrants in Vietnam, where 67% of IGRA-positive individuals accepted treatment and 88% of those completed treatment.^12^ Programmatically, overseas IGRA testing and LTBI treatment among refugees would produce additional pre-departure costs for the U.S. government, but would reduce costs for U.S. domestic TB programs through reduced costs of post-arrival testing and LTBI treatment, as well as from the potential reduction in TB cases and TB-related morbidity and mortality among refugees after U.S. arrival due to the preventive benefits of LTBI treatment. In this study, we compared the potential health impact, costs, and cost-effectiveness of three pre-departure IGRA testing and LTBI treatment strategies for U.S.-bound children and adult refugees from high-TB-burden countries.

## METHODS

### Study cohort

The population of interest was U.S.-bound refugees ≥2 years old, examined in a country with TB incidence ≥20 per 100,000. Country-of-examination was defined as the country where individuals received their overseas medical examination, which may differ from country-of-birth.

We created a synthetic study cohort of 100,000 individuals via simple random sampling with replacement from the dataset of refugee applicants registered in the CDC EDN system between 2013 and 2023 from countries-of-examination with a WHO-estimated TB incidence ≥20 per 100,000 in 2022.^13^ Data extracted for each individual included country-of-examination, country-of-birth, and age at U.S. entry. We imputed LTBI status for each individual in the study cohort by age and country-of-origin, based on estimates of LTBI prevalence among US-bound migrants from an earlier modeling study.^14^

### LTBI testing and treatment strategies

We evaluated three IGRA testing and LTBI treatment strategies for the study cohort:

*Strategy 1*: For refugees aged 2-14 years old, pre-departure IGRA testing is required. Individuals testing IGRA-positive receive a medical examination after U.S. arrival to identify TB disease, with LTBI treatment offered after TB has been ruled out. For refugees ≥15 years old, IGRA testing and LTBI treatment are offered after TB disease has been ruled out at the post-arrival medical examination. This strategy represents the pre-2024 screening requirements for U.S.-bound refugees testing in high-risk countries.

*Strategy 2*: Pre-departure IGRA testing is required for all refugees ≥2 years old. Individuals testing IGRA-positive are offered LTBI treatment after TB disease has been ruled out at the post-arrival medical examination. This strategy represents the current screening requirements for U.S.-bound refugees testing in high-risk countries, adopted in 2024.

*Strategy 3*: Pre-departure IGRA testing is required for all refugees ≥2 years old, with pre-departure LTBI treatment offered to individuals testing IGRA-positive, after TB disease has been ruled out (LTBI treatment is not required for U.S. entry). After U.S. arrival and domestic medical examination to rule out TB, LTBI treatment is offered again to those who decline LTBI treatment overseas. For those who initiate treatment overseas but do not complete the regimen, a proportion are assumed to re-initiate a full course of LTBI treatment during their post-arrival medical examination. We assumed that in some countries it may be infeasible to offer LTBI treatment to refugees before travel to the US, based on consultation with experts from the International Organization of Migration (IOM) (Text S1). For these countries we assumed LTBI treatment would only be offered after arrival (as in Strategy 2). In countries where pre-departure LTBI treatment was thought to be feasible (77 of 101 countries), we assumed it would be offered in refugee camps or healthcare settings accessible for refugees to receive directly observed therapy (DOT).

*‘No intervention’ scenario*: In addition to the three intervention strategies, we simulated a scenario that assumed no LTBI diagnosis or treatment would be offered before or after U.S. entry. Pre-departure IGRA testing would still be conducted for children as part of mandated TB disease screening. Incremental health outcomes and costs associated with each intervention strategy were calculated with respect to this ‘No Intervention’ scenario.

Under all strategies we assumed refugees would be screened for TB disease pre-departure, and those diagnosed with infectious TB disease required to complete TB treatment before departure for the United States, consistent with current U.S. Technical Instructions.^9^ We assumed QuantiFERON-TB Gold Plus would be used for IGRA testing, with 89% sensitivity and 98% specificity among individuals >14 years old, and 77% sensitivity and 96% specificity among individuals 2-14 years old.^15,16^ We assumed post-arrival examination would occur one month after U.S. arrival. Figure S1 illustrates each intervention strategy.

For LTBI treatment initiated overseas, we assumed once-weekly isoniazid and rifapentine for 3 months (3HP) by DOT would be used. This regimen was selected in consultation with IOM experts based on its short treatment course compared to other regimens, while not involving the daily dosing of shorter regimens. We assumed treatment was via DOT (vs. self-administered) given likely higher adherence and completion in the context of overseas refugee programs. We assumed 65% of individuals offered LTBI treatment overseas would initiate treatment, and of those initiating treatment 88% would complete the regimen (Table 1).^12^ For LTBI treatment initiated in the United States, we assumed 3HP would be prescribed, via self-administered therapy (SAT). While both 4R (4 months of daily rifampin) and 3HP are popular LTBI treatment regimens in the United States,^17^ we chose 3HP because of the high cost of the 4R regimen at the time of this study.^18^ We assumed 88% of individuals would attend the post-arrival refugee medical examination, 76% of individuals offered LTBI treatment after U.S. entry would initiate the regimen, and of these individuals 63% would complete treatment, based on CDC EDN data.^11^ For all strategies, we assumed LTBI treatment would only be offered after TB disease was ruled out. If diagnosed with TB disease, a standard first-line treatment would be prescribed.^19^ Key parameter values are reported in Table 1, with supplementary details in Tables S2-S3.

**Table 1.** Key parameters for IGRA testing and LTBI treatment strategies.

| Parameter definitions | Point estimate<br>(95% interval) | Source |
| --- | --- | --- |
| **Test performance parameters** |  |  |
| Specificity of IGRA for LTBI (%) |  |  |
| *Aged < 15 years* | 96 (84, 97) | 16 |
| *Aged ≥ 15 years* | 98 (95, 99) | 15 |
| Sensitivity of IGRA for LTBI (%) |  |  |
| *Aged < 15 years* | 77 (43, 94) | 16 |
| *Aged ≥ 15 years* | 89 (84, 94) | 15 |
| Specificity of TB diagnosis (%) | 100 | Assumption |
| Sensitivity of TB diagnosis (%) | 100 | Assumption |
| **Overseas IGRA testing and LTBI treatment (when implemented, see Figure S1)** |  |  |
| Percentage (%) screened with IGRA (children: Strategies 1-3, adults: Strategies 2-3) | 100 | Assumption |
| Percentage (%) initiated LTBI treatment (3HP), among those with a positive IGRA result (children: Strategy 3, adults: Strategy 3) | 65 (51, 79) | 12 |
| Percentage (%) completed the full course of 3HP, or completed a sufficient proportion of 3HP prior to U.S. entry such that a re-initiation is not required, among those initiating 3HP | 88 (83, 92) | 12 |
| **Domestic IGRA testing and LTBI treatment (when implemented, see Figure S1)** |  |  |
| Percentage (%) attended domestic medical evaluation for refugees | 88 (83, 92) | Assumption |
| Percentage (%) screened with IGRA, conditional on not tested with IGRA overseas and no signs of TB disease | 100 | Assumption |
| Percentage (%) offered LTBI treatment (3HP), conditional on a positive IGRA result from overseas or domestic testing, with TB disease ruled out and one of the following: not offered 3HP overseas, offered but did not initiate 3HP overseas, or initiated but did not complete a sufficient proportion of 3HP signaling a need to re-initiate treatment | 100 | Assumption |
| Percentage (%) initiated 3HP, conditional on being offered 3HP, regardless of LTBI treatment history | 76 (69, 82) | Electronic Disease Notification* |
| Percentage (%) completed 3HP, conditional on having initiated 3HP, regardless of LTBI treatment history | 63 (56, 69) | Electronic Disease Notification* |

Abbreviations: IGRA, interferon gamma release assay; LTBI, latent TB infection; 3HP, once-weekly isoniazid and rifapentine for 3 months. Parameter values apply to all strategies unless indicated. The full list of model parameters is presented in Table S2.
\* Based on Electronic Disease Notification data for 2013-2023 (only data for children 2-14 years were used to inform these parameters because of poor quality of the adults data).<sup>11</sup>

### Discrete event simulation model

We adapted an existing individual-based discrete event simulation model to project TB-related events for individuals in the study cohort, over a lifetime horizon.^20^ For each individual, the model simulated future TB incidence for the baseline scenario based on incidence rates reported for U.S. resident migrants,^21^ by age, years since U.S. entry, and country-of-birth. Under Strategies 1-3, we assumed the provision of LTBI treatment would reduce future TB incidence in the modelled population, as determined by rates of regimen initiation, completion, and efficacy (Tables 1, S2-3).^22^ Individuals developing TB disease were assumed to receive TB diagnosis and treatment. Background mortality rates derived from U.S. life tables reported for non-U.S.-born individuals,^23^ and TB-associated mortality rates before and during treatment, were derived from national TB outcomes data.^24^ Model parameters are reported in Tables 1 and S2-3. The model was programmed in R version 4.2.3.^25^

### Health outcomes, costs, and economic evaluation

We estimated the number of TB cases averted and quality adjusted life years (QALYs) gained over the lifetime of each individual in the study cohort, based on differences in simulated TB incidence and mortality between scenarios, and published utility weights (Table S2). In the main analysis, for pre-departure IGRA testing and LTBI treatment, we assumed the mean unit cost of IGRA was $50 (based on communications with experts from IOM), and the mean unit cost of a full course of 3HP was $214 for adults and $208 for children (calculation details in Tables S4-1, S4-2). For IGRA testing and LTBI treatment in the United States, we assumed the unit cost for IGRA was $62,^26^ and the mean unit cost of 3HP was $486 for adults and $528 for children (Tables S4-1, S4-2). All costs were converted to 2023 US dollars. We conducted a cost-effectiveness analysis from the TB services perspective, with future QALYs and costs discounted at 3% annually.

### Sensitivity and scenario analyses

We conducted a probabilistic sensitivity analysis to calculate measures of uncertainty (equal-tailed 95% uncertainty intervals) around study outcomes. Table S3 reports parameter distributions. Additionally, we conducted one-way sensitivity analyses for several key parameters: the proportion of refugees with a positive IGRA result who successfully complete LTBI treatment overseas; the proportion of refugees who attend domestic medical follow-up medical examination after having initiated LTBI treatment overseas; and the unit cost of 3HP overseas. We also conducted scenario analyses by varying the LTBI prevalence of the refugee populations to 50% and 200% of the base-case value.

## RESULTS

Among the study cohort, 33% were children aged 2-14 (n = 33,191) with a mean entry age of 8 years (standard deviation = 4). The mean entry age of the adult population (n = 66,809) was 32 years (std dev = 14). Table S1 reports countries-of-examination and countries-of-birth represented in the study cohort. Average LTBI prevalence was estimated as 9.6% for children and 28.0% for adults in the study cohort.

### Process and health outcomes

Table 2 reports the percentage of child and adult refugee populations retained in the pre-departure and post-arrival testing and treatment care cascade under each strategy. For both children and adults, the fraction of refugees completing LTBI treatment was higher in Strategy 3 compared to Strategies 1-2 (7.1% versus 4.5% for children; 19.3% versus 10.9% for adults).

**Table 2.** Percentage of the simulated refugee cohort tested with IGRA and received LTBI treatment, overseas and in the United States.

|  | Overseas services<br>(% of study cohort by age group, 95% UI) |  |  | U.S. services<br>(% of study cohort by age group, 95% UI) |  |  | Total services completed |  |
| --- | --- | --- | --- | --- | --- | --- | --- | --- |
|  | IGRA positive | Initiated 3HP | Completed 3HP | IGRA positive | Initiated 3HP | Completed 3HP | % of study cohort by age group (95% UI) | % of IGRA-positives, by age group (95% UI) |
| **Children (n = 33,191)** |  |  |  |  |  |  |  |  |
| Strategy 1, 2* | 10.7<br>(6.1, 18.1) | .. | .. | .. | 7.1<br>(4.0, 12.1) | 4.5<br>(2.5, 7.6) | 4.5<br>(2.5, 7.6) | 41.7<br>(36.1, 47.6) |
| Strategy 3 | 10.7<br>(6.1, 18.1) | 5.1<br>(3.1, 7.2) | 4.8<br>(2.6, 6.7) | .. | 3.7<br>(1.5, 8.0) | 2.3<br>(0.9, 5.0) | 7.1<br>(4.3, 10.8) | 67.2<br>(55.7, 78.3) |
| **Adults (n = 66,809)** |  |  |  |  |  |  |  |  |
| Strategy 1 | .. | .. | .. | 23.0<br>(21.0, 25.2) | 17.3<br>(15.2, 19.6) | 10.9<br>(9.2, 12.7) | 10.9<br>(9.2, 12.7) | 47.4<br>(41.3, 53.6) |
| Strategy 2 | 26.1<br>(24.2, 28.3) | .. | .. | .. | 17.3<br>(15.2, 19.6) | 10.9<br>(9.2, 12.7) | 10.9<br>(9.2, 12.7) | 41.7<br>(35.9, 47.8) |
| Strategy 3 | 26.1<br>(24.2, 28.3) | 16.8<br>(13.0, 20.7) | 14.9<br>(11.4, 18.5) | .. | 7.0<br>(4.6, 9.4) | 4.5<br>(2.6, 6.7) | 19.3<br>(16.8, 22.0) | 73.9<br>(65.8, 81.7) |

Abbreviations: IGRA, interferon gamma release assay; LTBI, latent TB infection; UI, uncertainty interval; %, percentage; 3HP, once-weekly isoniazid and rifapentine for 3 months; .., not applicable. Results are based on simulation of a synthetic cohort based on the demographics of refugees in the CDC Electronic Disease Notification data between 2013-2023.
\* Strategy 1: pre-departure IGRA testing for children (2-14 years) and post-arrival IGRA testing for adults (>14 years), with 3HP offered in the United States for IGRA-positive individuals after ruling out TB disease. Strategy 2: pre-departure IGRA testing children and adults, and post-arrival 3HP offered for IGRA-positive individuals after ruling out TB disease. Strategy 3: pre-departure IGRA testing for children and adults, and pre-departure 3HP offered for IGRA-positive individuals testing after ruling out TB disease, with 3HP re-offered in the United States for individuals not completing treatment before U.S. arrival.

Without any LTBI treatment, we estimated that 54 (95% uncertainty interval: 40–69) out of 33,191 children and 409 (365–466) out of 66,809 adults in the modeled refugee cohort would develop TB disease during their lifetime in the United States. For both children and adults in the study cohort, Strategy 3 was estimated to result in the greatest reduction in future TB incidence (Table 3). Lifetime TB incidence in Strategy 3 was estimated at 26 (15–41) cases among children (a 52% (29–73%) reduction compared to the ‘No Intervention’ scenario) and 165 (123–210) cases among adults (a 60% (51–69%) reduction compared to the ‘No Intervention’ scenario).

**Table 3.** Health and cost outcomes for the simulated refugee cohort.

| Strategy | Total TB cases in the U.S.<br>(n, 95% UI) | Reduction in U.S. TB cases, relative to the No intervention scenario*<br>(%, 95% UI) | Per-person IGRA testing and 3HP costs overseas<br>(\$ per refugee, 95% UI) | Per-person IGRA testing and 3HP costs in the U.S.<br>(\$ per refugee, 95% UI) | Per-person IGRA testing and 3HP costs in the U.S.<br>(\$ per refugee, 95% UI) | Total per-person cost (\$ per refugee, 95% UI) |
| --- | --- | --- | --- | --- | --- | --- |
| **Children (n = 33,191)** |  |  |  |  |  |  |
| *No intervention scenario\** | 54<br>(40 – 69) | .. | .. | .. | 25.4<br>(14.7 – 39.7) | 25.4<br>(14.7 – 39.7) |
| *Strategy 1, 2* | 38<br>(24 – 52) | 48.1<br>(8.8 – 57.2) | 0.0<br>(0.0 – 0.0)** | 37.2<br>(17.0 – 72.3) | 21.6<br>(13.3 – 34.0) | 59.0<br>(36.2 – 97.1) |
| *Strategy 3* | 26<br>(15 – 41) | 51.9<br>(29.0 – 73.3) | 10.1<br>(4.04 – 19.0) | 19.4<br>(6.8 – 45.7) | 13.5<br>(6.8 – 23.7) | 43.1<br>(25.7 – 75.0) |
| **Adults (n = 66,809)** |  |  |  |  |  |  |
| *No intervention scenario* | 409<br>(365 – 466) | ..<br>(0.0 – 0.0) | .. | .. | 107.9<br>(68.3 – 158.0) | 107.9<br>(68.3 – 158.0) |
| *Strategy 1* | 268<br>(226 – 317) | 34.4<br>(27.9 – 41.5) | 0.0<br>(0.0 – 0.0) | 138.7<br>(98.7 – 189.4) | 82.3<br>(54.6 – 118.7) | 221.9<br>(170.9 – 280.0) |
| *Strategy 2* | 268<br>(226 – 317) | 34.5<br>(28.0 – 41.5) | 50.0<br>(28.8 – 76.9) | 84.0<br>(46.5 – 133.6) | 82.1<br>(54.6 – 118.7) | 216.2<br>(163.6 – 277.8) |
| *Strategy 3* | 165<br>(123 – 210) | 59.8<br>(50.8 – 68.6) | 82.8<br>(53.0 – 117.4) | 33.9<br>(16.4 – 57.6) | 47.9<br>(29.2 – 73.3) | 164.6<br>(126.2 – 212.1) |

Abbreviations: IGRA, interferon gamma release assay; UI, uncertainty interval; 3HP, once-weekly isoniazid and rifapentine for 3 months; .., not applicable. Results are based on simulation of a synthetic cohort based on the demographics of refugees in the CDC Electronic Disease Notification data between 2013-2023. All costs represent discounted 2023 US dollars.
\* No intervention scenario: No IGRA testing and LTBI treatment for the refugee cohort, except for IGRA testing as part of mandatory TB screening in children. Strategy 1: pre-departure IGRA testing for children (2-14 years) and post-arrival IGRA testing for adults (>14 years), with 3HP offered in the United States for IGRA-positive individuals after ruling out TB disease Strategy 2: pre-departure IGRA testing children and adults, and post-arrival 3HP offered for IGRA-positive individuals after ruling out TB disease; Strategy 3: pre-departure IGRA testing for children and adults, and pre-departure 3HP offered for IGRA-positive individuals testing after ruling out TB disease, with 3HP re-offered in the United States for individuals not completing treatment before U.S. arrival.
\*\* IGRA testing and treatment costs overseas are zero in Strategies 1-2 for children because IGRA testing is part of mandated TB disease screening for children. IGRA costs are included for adults as this testing is undertaken for LTBI diagnosis.

For children, compared to the ‘No Intervention’ scenario, IGRA testing and LTBI treatment provided through Strategies 1-2 was estimated to produce a gain of 0.78 (0.24–1.72) undiscounted QALYs per 1000 refugees, while for Strategy 3 there were 1.37 (0.57–2.45) undiscounted QALYs gained per 1000 refugees. For adults, the number of undiscounted QALYs gained per 1000 refugees were similar in Strategy 1 (3.85, 2,80–5.17) and Strategy 2 (3.82, 2.76–5.08), and were higher in Strategy 3 (6.48, 4.93–8.33).

### Cost outcomes and cost-effectiveness analysis

For both children and adults, the estimated total costs associated with Strategy 3 were lower than the other two strategies. For Strategy 3, total costs (summing costs incurred overseas and within the United States) were $43 (26–75) per child, compared to $59 (36–97) per child for Strategies 1 and 2. For adults, per-person costs were $222 (171–280), $216 (164–278), and $165 (126–212) for Strategies 1, 2, and 3, respectively. As compared to Strategies 1-2, we estimated that Strategy 3 would produce greater costs for pre-departure LTBI treatment and reduced costs for post-arrival LTBI testing and treatment, as well as reductions in the cost of treating TB disease over their lifetime in the United States (Table 3).

Strategy 3 (IGRA testing and LTBI treatment offered overseas) produced greater health gains and lower incremental costs compared to Strategies 1 and 2 (Table 4), and therefore ‘dominated’ these two other strategies. As compared to the ‘No Intervention’ scenario, Strategy 3 was estimated to have an incremental cost-effectiveness ratio (ICER) of US $45,000 per QALY gained for children, and $21,111 per QALY gained for adults.

**Table 4.** Cost-effectiveness of alternative strategies for IGRA testing and LTBI treatment among the simulated refugee cohort.

| Strategy | Total QALY<br>(per refugee, 95% UI) | Total cost<br>(per refugee, 95% UI) | Incremental<br>QALY | Incremental<br>cost | ICER<br>(\$ / QALY<br>gained) |
| --- | --- | --- | --- | --- | --- |
| **Children** |  |  |  |  |  |
| *No intervention scenario\** | 29.8816<br>(29.8811 – 29.8818) | 25<br>(16 – 37) | .. | .. | .. |
| *Strategy 1, 2* | 29.8818<br>(29.8815 – 29.8820) | 59<br>(30– 104) | .. | .. | Dominated by Strategy 3** |
| *Strategy 3* | 29.8820<br>(29.8817 – 29.8821) | 43<br>(17 – 88) | 0.0004<br>(0.0002 – 0.0008) | 18<br>(-1 – 48) | 45,000 |
| **Adults** |  |  |  |  |  |
| *No intervention scenario* | 25.5178<br>(25.5168 – 25.5187) | 108<br>(79 – 143) | .. | .. | .. |
| *Strategy 1* | 25.5194<br>(25.5187 – 25.5201) | 203<br>(114 – 307) | .. | .. | Dominated by Strategy 3 |
| *Strategy 2* | 25.5194<br>(25.5186 – 25.5200) | 216<br>(132 – 327) | .. | .. | Dominated by Strategy 3 |
| *Strategy 3* | 25.5205<br>(25.5199 – 25.5210) | 165<br>(101 – 246) | 0.0027<br>(0.0020 – 0.0035) | 57<br>(10 – 104) | 21,111 |

Abbreviation: ICER, incremental cost-effectiveness ratio; QALY, quality-adjusted life year; UI, uncertainty interval; IGRA, interferon gamma release assay; LTBI, latent TB infection; 3HP, once-weekly isoniazid and rifapentine for 3 months; .., not applicable. Costs (2023 USD) and QALYs discounted at 3%. Costs assessed from a TB services perspective (overseas TB screening not included) with a lifetime analytic horizon.
\* No intervention scenario: No IGRA testing and LTBI treatment for the refugee cohort, except for IGRA testing as part of mandatory TB screening in children.
Strategy 1: pre-departure IGRA testing for children (2-14 years) and post-arrival IGRA testing for adults (>14 years), with 3HP offered in the United States for IGRA-positive individuals after ruling out TB disease; Strategy 2: pre-departure IGRA testing children and adults, and post-arrival 3HP offered for IGRA-positive individuals after ruling out TB disease; Strategy 3: pre-departure IGRA testing for children and adults, and pre-departure 3HP offered for IGRA-positive individuals testing after ruling out TB disease, with 3HP re-offered in the United States for individuals not completing treatment before U.S. arrival.
\*\* 'Dominated' indicates that the strategy produces worse health outcomes and has higher costs than another strategy (or combination of strategies) included in the analysis.

### Sensitivity and scenario analyses

In one-way sensitivity analyses, Strategy 3 was estimated to result in higher QALYs gained and lower additional costs, as compared to Strategy 2, across plausible values of the treatment completion rate for overseas LTBI treatment, post-arrival follow-up rate among those who initiated LTBI treatment overseas, and the cost of 3HP overseas, except when the treatment completion rate was close to 2% or when the cost of LTBI treatment overseas was greater than $550 for adults and $535 for children (Table 5).

**Table 5.**
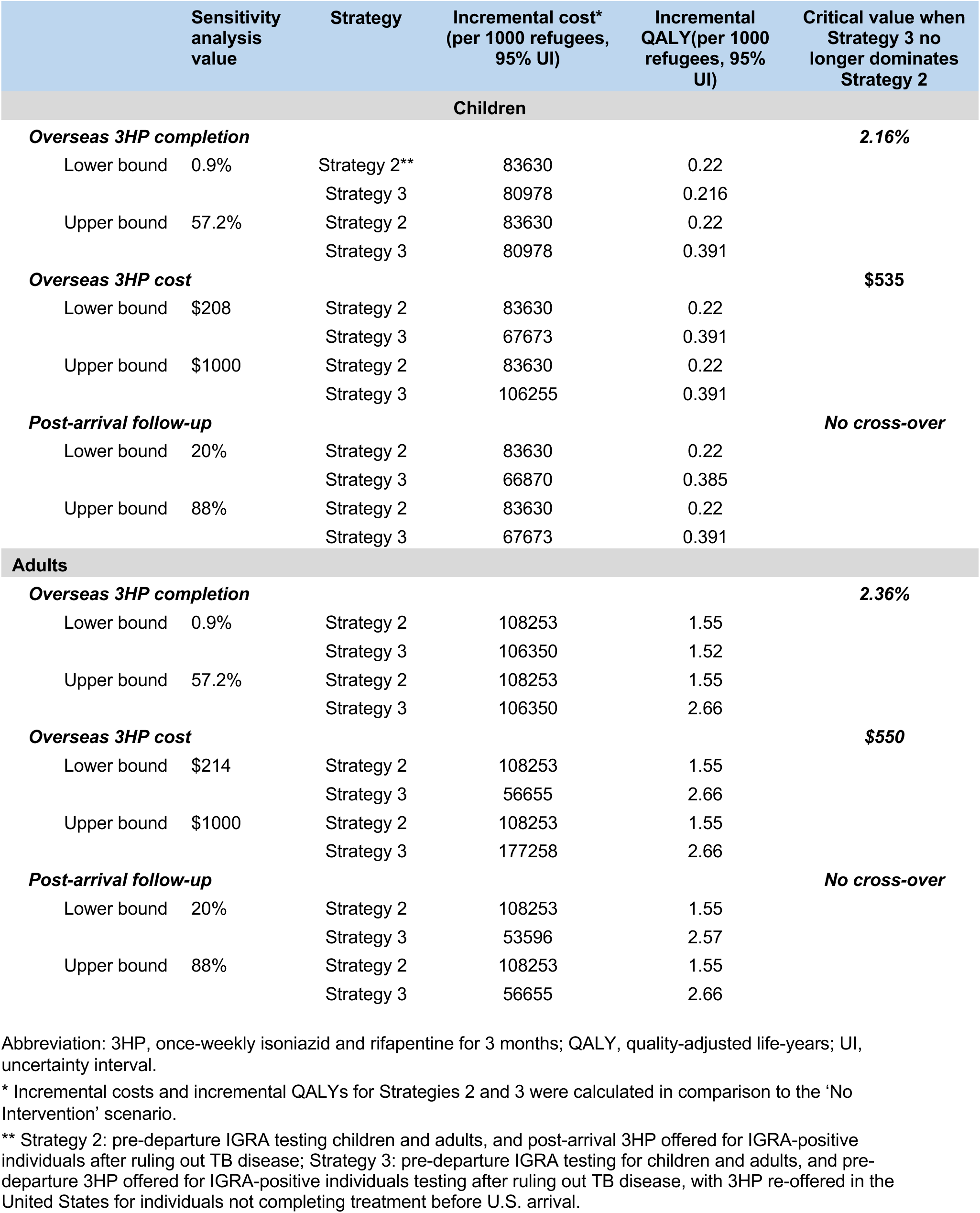
One-way sensitivity analyses varying key model parameters.

|  | Sensitivity analysis value | Strategy | Incremental cost* (per 1000 refugees, 95% UI) | Incremental QALY(per 1000 refugees, 95% UI) | Critical value when Strategy 3 no longer dominates Strategy 2 |
| --- | --- | --- | --- | --- | --- |
| **Children** |  |  |  |  |  |
| **Overseas 3HP completion** |  |  |  |  | **2.16%** |
| Lower bound | 0.9% | Strategy 2** | 83630 | 0.22 |  |
|  |  | Strategy 3 | 80978 | 0.216 |  |
| Upper bound | 57.2% | Strategy 2 | 83630 | 0.22 |  |
|  |  | Strategy 3 | 80978 | 0.391 |  |
| **Overseas 3HP cost** | | | | | **\$535** |
| Lower bound | \$208 | Strategy 2 | 83630 | 0.22 | |
|  |  | Strategy 3 | 67673 | 0.391 |  |
| Upper bound | \$1000 | Strategy 2 | 83630 | 0.22 | |
|  |  | Strategy 3 | 106255 | 0.391 |  |
| **Post-arrival follow-up** |  |  |  |  | **No cross-over** |
| Lower bound | 20% | Strategy 2 | 83630 | 0.22 |  |
|  |  | Strategy 3 | 66870 | 0.385 |  |
| Upper bound | 88% | Strategy 2 | 83630 | 0.22 |  |
|  |  | Strategy 3 | 67673 | 0.391 |  |
| **Adults** |  |  |  |  |  |
| **Overseas 3HP completion** |  |  |  |  | **2.36%** |
| Lower bound | 0.9% | Strategy 2 | 108253 | 1.55 |  |
|  |  | Strategy 3 | 106350 | 1.52 |  |
| Upper bound | 57.2% | Strategy 2 | 108253 | 1.55 |  |
|  |  | Strategy 3 | 106350 | 2.66 |  |
| **Overseas 3HP cost** | | | | | **\$550** |
| Lower bound | \$214 | Strategy 2 | 108253 | 1.55 | |
|  |  | Strategy 3 | 56655 | 2.66 |  |
| Upper bound | \$1000 | Strategy 2 | 108253 | 1.55 | |
|  |  | Strategy 3 | 177258 | 2.66 |  |
| **Post-arrival follow-up** |  |  |  |  | **No cross-over** |
| Lower bound | 20% | Strategy 2 | 108253 | 1.55 |  |
|  |  | Strategy 3 | 53596 | 2.57 |  |
| Upper bound | 88% | Strategy 2 | 108253 | 1.55 |  |
|  |  | Strategy 3 | 56655 | 2.66 |  |

Abbreviation: 3HP, once-weekly isoniazid and rifapentine for 3 months; QALY, quality-adjusted life-years; UI, uncertainty interval.
\* Incremental costs and incremental QALYs for Strategies 2 and 3 were calculated in comparison to the 'No Intervention' scenario.
\*\* Strategy 2: pre-departure IGRA testing children and adults, and post-arrival 3HP offered for IGRA-positive individuals after ruling out TB disease; Strategy 3: pre-departure IGRA testing for children and adults, and pre-departure 3HP offered for IGRA-positive individuals testing after ruling out TB disease, with 3HP re-offered in the United States for individuals not completing treatment before U.S. arrival.

Strategy 3 remained the dominant strategy in sensitivity analyses in which we estimated results with LTBI prevalence rates at both double and half the values used in the primary analysis (Tables S5-1, S5-2).

## DISCUSSION

This analysis examined the potential health impact and cost-effectiveness of alternative approaches for diagnosing and treating LTBI among refugees resettling to the United States from high-TB-burden countries. We found that a strategy requiring pre-departure IGRA testing and offering pre-departure LTBI treatment in the country-of-examination could result in greater treatment initiation and completion—and therefore reduce future TB incidence—relative to current approaches that offer LTBI treatment only after U.S. entry. This novel approach was also estimated to reduce TB-associated costs, particularly for US-based health services, and was cost-effective compared to existing approaches. When LTBI treatment was offered both before departure and after arrival (Strategy 3), this increased the number of opportunities for refugees to initiate and complete treatment, contributing to higher overall LTBI treatment completion and greater health benefits.

Several prior studies have examined the cost-effectiveness of LTBI testing and treatment for migrants residing in low incidence high-income countries,^27–29^ including several conducted in the United States.^30–33^ These studies have found intervention health benefits and cost-effectiveness to vary by setting, target population, and intervention design, with greater health benefits and more favorable cost-effectiveness ratios estimated when interventions are offered to individuals from high-TB-burden countries or to populations with additional TB risk factors. A recent feasibility study among U.S.-bound immigrants in Vietnam found that almost half of visa applicants agreed to receive voluntary IGRA testing. Among those testing positive, most initiated (67%), and completed initiated LTBI treatment (88%).^12^ Refugees represent a population at high risk of TB infection and disease due to higher prior TB exposure compared to other migrant populations.^34,35^ Several studies have investigated LTBI testing and treatment for refugee and asylee populations.^36–38^ Of these, one investigated the potential for LTBI testing (via tuberculin skin test) and treatment before U.S. entry as compared to post-arrival testing and treatment.^38^ It found that pre-departure LTBI testing and treatment could produce cost-savings and health benefits compared to a post-arrival intervention, because of greater losses along the care cascade for a post-arrival intervention compared to overseas interventions. Similarly, a Canadian study of pre-departure IGRA testing and post-arrival LTBI treatment found this intervention to be cost-effective for migrants with high prior TB exposure.^39^ A study conducted in Australia found that migrant LTBI testing and treatment would be substantially more impactful with testing conducted before versus after arrival.^40^ While the present study focused on an additional intervention that was not widely considered in these earlier studies (the addition of pre-departure LTBI treatment to pre-departure testing and post-arrival LTBI treatment), the conclusions are consistent with these earlier results, with greater effectiveness and cost-effectiveness estimated for interventions that increase the proportion of eligible individuals that complete the LTBI treatment regimen, by making services available earlier in the migration process. Also, consistent with an earlier study,^28^ our analysis found lower health benefits for children receiving LTBI screening and treatment as compared to adults. In our analysis, the lower estimate of QALYs gained (and higher cost-effectiveness ratios) for children vs. adults resulted from lower LTBI prevalence among younger age groups, reducing the number of future TB cases that could be averted.

In our study, as in earlier studies, losses along the care cascade substantially reduced intervention effectiveness. Lack of access and engagement with healthcare services, non-acceptance of testing, failure to initiate treatment, and discontinuation of treatment before completion are all challenges for TB programs globally. The context of pre-departure LTBI treatment may result in higher intervention acceptance and adherence among refugees than in other settings, but efforts to facilitate treatment completion will still be important.

As compared to the other intervention strategies, we found the strategy with pre-departure LTBI treatment (Strategy 3) to have the lowest total costs. This resulted from a substantial reduction in costs incurred within the United States, which outweighed the added expenses of the additional services provided overseas. As overseas refugee services (funded by the Department of State) are supported by a different budget than domestic services, a decision to implement Strategy 3 (due to its lower overall costs and greater health benefits compared to other intervention strategies), would require additional funding to be allocated to support the new services provided through overseas refugee programs.

In addition to health benefits and cost-effectiveness, health programs must consider acceptability, ethics, and feasibility when considering new interventions. While we did not examine these aspects explicitly, intervention strategies were designed with these concerns in mind. Under all scenarios, we assumed that individuals receiving an LTBI diagnosis would be recommended to receive LTBI treatment, but treatment would not be required for U.S. entry (in contrast to infectious TB disease treatment). While making LTBI treatment mandatory would raise treatment completion rates, it was considered unduly coercive given the magnitude of the health risks conferred by LTBI (substantially less than for TB disease), and hence not modeled in this study.^41,42^ Feasibility was a consideration in restricting Strategy 3 (pre-departure LTBI treatment) to a subset of countries. While a pilot study has demonstrated the feasibility of pre-departure voluntary LTBI treatment for U.S. immigrant visa applicants,^12^ the context of refugee applicants is sufficiently different from other migrants, such that empirical confirmation would be required before any change in intervention strategy.

This study has several limitations. First, future health outcomes were estimated using mathematical modeling, rather from empirical data. This approach is shared by the majority of cost-effectiveness studies of LTBI interventions, given that the TB incidence reductions produced by LTBI treatment require large sample sizes for accurate measurement, and will be spread over many years, making observational studies difficult. However, empirical evidence on migrant LTBI interventions is consistent with the modelling results, with a follow-up study of IGRA-positive migrants accepting LTBI treatment in England finding a hazard ratio of 0.14 (95%CI: 0.06–0.32) for future TB as compared to IGRA-positive migrants who did not receive treatment.^43^ Moreover, we used evidence on TB incidence among all migrants entering the United States (describing TB risks with increasing time since U.S. entry), and it is possible these estimates may vary for refugees.^21^ Second, while scenarios were designed and parameter values chosen in consultation with individuals experienced with refugee programs, this expert input may not anticipate all challenges involved with a novel approach. Similarly, the costs for overseas LTBI treatment remain highly uncertain and are likely to vary across countries. There is also uncertainty around the costs for LTBI treatment in the United States, exemplified by the 1200% jump in the price of rifampin over 2020-2023.^44^ ^18^ Lastly, we did not consider secondary TB transmission, and only calculated health outcomes for individuals receiving the intervention. While including secondary transmission would have produced greater estimated health benefits, empirical data suggest that onward transmission in the United States is rare,^45^ and hence the number of secondary TB cases would be low.

In many settings, recently arrived refugees have some of the highest risks for incident TB disease. Therefore, identifying interventions that can reduce TB risk among the refugee population—in ways that are cost-effective, feasible, and respectful of individual autonomy—is a high public health priority. This study found that pre-departure IGRA testing and voluntary LTBI treatment would be cost-effective compared to current prevention approaches, reducing TB risk for a traditionally underserved population at high risk of TB disease.

## Data Availability

Model code input parameters, and outputs available upon request.

## Acknowledgements

This project was funded by the U.S. Centers for Disease Control and Prevention, National Center for HIV/AIDS, Viral Hepatitis, STD, and TB Prevention Epidemiologic and Economic Modeling Agreement (#5NU38PS004644). We thank Amera Khan for sharing summary statistics for findings from the study “Khan A, *et al*. Overseas Treatment of Latent Tuberculosis Infection in US–Bound Immigrants. *Emerg Infect Dis* 2022;28(3):582-590”; Andrew Hill for sharing the detailed results of the TB risk model reported in the study “Hill AN, *et al*. High-resolution estimates of tuberculosis incidence among non-U.S.-born persons residing in the United States, 2000–2016. *Epidemics* 2020; **33**: 100419”; and Alexander Klosovsky, Salma Taher, Farah Amin, and Damaris Miriti from the International Organization of Migration for their input on our model input parameters related to IGRA testing and LTBI treatment overseas for U.S.-bound refugees. Additionally, we thank the following for their valuable input to our manuscript: Carla Winston, U.S. CDC; C Robert Horsburgh Jr, Departments of Global Health, Epidemiology, Biostatistics, and Medicine, Boston University; and Joshua Salomon, Department of Health Policy, Stanford University School of Medicine. Lastly, we thank Teresa Puente and Stephanie Su for administrative support. The findings and conclusions in this paper are those of the authors and do not represent the view of the funding sources or the authors’ affiliated institutions.

## REFERENCES

1. Pareek, M., Greenaway, C., Noori, T., Munoz, J. & Zenner, D. The impact of migration on tuberculosis epidemiology and control in high-income countries: a review. BMC Med. 14, 48 (2016).

2. OTIS TB Data 1993-2023 Request. https://wonder.cdc.gov/TB-v2023.html.

3. Menzies, N. A., Hill, A. N., Cohen, T. & Salomon, J. A. The impact of migration on tuberculosis in the United States. Int. J. Tuberc. Lung Dis. Off. J. Int. Union Tuberc. Lung Dis. 22, 1392–1403 (2018).

4. US Preventive Services Task Force. Screening for Latent Tuberculosis Infection in Adults: US Preventive Services Task Force Recommendation Statement. JAMA 329, 1487–1494 (2023).

5. Sterling, T. R. Guidelines for the Treatment of Latent Tuberculosis Infection: Recommendations from the National Tuberculosis Controllers Association and CDC, 2020. MMWR Recomm. Rep. 69, (2020).

6. Tuberculosis prevention and care among refugees and other populations in humanitarian settings: an interagency field guide. https://www.who.int/publications/i/item/9789240042087.

7. Innovative solutions for the elimination of tuberculosis among migrants and refugees. https://www.who.int/publications/m/item/innovative-solutions-for-the-elimination-of-tuberculosis-among-migrants-and-refugees.

8. Immigration and Nationality Act | USCIS. https://www.uscis.gov/laws-and-policy/legislation/immigration-and-nationality-act (2019).

9. Tuberculosis Technical Instructions for Panel Physicians | CDC. https://www.cdc.gov/immigrantrefugeehealth/panel-physicians/tuberculosis.html?CDC_AA_refVal=https%3A%2F%2Fwww.cdc.gov%2Fimmigrantrefugeehealth%2Fexams%2Fti%2Fpanel%2Ftuberculosis-panel-technical-instructions.html#indicators.

10. CDC. Refugee Health Domestic Guidance. Immigrant and Refugee Health https://www.cdc.gov/immigrant-refugee-health/hcp/domestic-guidance/index.html (2024).

11. Unpublished CDC data from the Electronic Disease Notification 2013-2023 (only data children 2-14 years, provided by Christina Phares, Division of Global Migration Health, 2024-03-20.

12. Khan, A. et al. Overseas Treatment of Latent Tuberculosis Infection in US–Bound Immigrants. Emerg. Infect. Dis. 28, 582–590 (2022).

13. Global Tuberculosis Report 2023. https://www.who.int/teams/global-tuberculosis-programme/tb-reports/global-tuberculosis-report-2023.

14. Menzies, N. A. et al. The long-term effects of domestic and international tuberculosis service improvements on tuberculosis trends within the USA: a mathematical modelling study. Lancet Public Health 9, e573–e582 (2024).

15. Jonas, D. E. et al. Screening for Latent Tuberculosis Infection in Adults: Updated Evidence Report and Systematic Review for the US Preventive Services Task Force. JAMA 329, 1495–1509 (2023).

16. Hirabayashi, R., Nakayama, H., Yahaba, M., Yamanashi, H. & Kawasaki, T. Utility of interferon-gamma releasing assay for the diagnosis of active tuberculosis in children: A systematic review and meta-analysis. J. Infect. Chemother. 30, 516–525 (2024).

17. Feng, P.-J. I., Horne, D. J., Wortham, J. M. & Katz, D. J. Trends in tuberculosis clinicians’ adoption of short-course regimens for latent tuberculosis infection. J. Clin. Tuberc. Mycobact. Dis. 33, 100382 (2023).

18. Logistics, O. of P., Acquisition and. VA.gov | Veterans Affairs. https://www.va.gov/opal/nac/fss/pharmprices.asp.

19. Saukkonen, J. J. et al. Updates on the Treatment of Drug-Susceptible and Drug-Resistant Tuberculosis: An Official ATS/CDC/ERS/IDSA Clinical Practice Guideline. Am. J. Respir. Crit. Care Med. 211, 15–33 (2025).

20. Hsieh, Y. L. et al. Cost-effectiveness of screening with transcriptional signatures for incipient TB among U.S. migrants. PLOS Med. 22, e1004603 (2025).

21. Hill, A. N., Cohen, T., Salomon, J. A. & Menzies, N. A. High-resolution estimates of tuberculosis incidence among non-U.S.-born persons residing in the United States, 2000–2016. Epidemics 33, 100419 (2020).

22. Sterling, T. R. et al. Three Months of Rifapentine and Isoniazid for Latent Tuberculosis Infection. N. Engl. J. Med. 365, 2155–2166 (2011).

23. Lauren Medina, Shannon Sabo, and Jonathan Vespa. Living Longer: Historical and Projected Life Expectancy in the United States, 1960 to 2060. https://www.census.gov/content/dam/Census/library/publications/2020/demo/p25-1145.pdf (2020).

24. Unpublished 2022 CDC data for non-US born population residing in the United States for less than 10 years, provided by Julie Self, Lauren Lambert, and Bob Pratt from the U.S. CDC Surveillance Team, Division of Tuberculosis Elimination, 2022-10-06.

25. R Core Team (2023). R: A language and environment for statistical computing. R. Foundation for Statistical Computing.

26. Clinical Laboratory Fee Schedule | CMS. https://www.cms.gov/medicare/medicare-fee-for-service-payment/clinicallabfeesched.

27. Greenaway, C. et al. The effectiveness and cost-effectiveness of screening for latent tuberculosis among migrants in the EU/EEA: a systematic review. Euro Surveill. Bull. Eur. Sur Mal. Transm. Eur. Commun. Dis. Bull. 23, 17–00543 (2018).

28. Zammarchi, L. et al. A scoping review of cost-effectiveness of screening and treatment for latent tuberculosis infection in migrants from high-incidence countries. BMC Health Serv. Res. 15, 412 (2015).

29. Zenner, D., Hafezi, H., Potter, J., Capone, S. & Matteelli, A. Effectiveness and cost-effectiveness of screening migrants for active tuberculosis and latent tuberculous infection. Int. J. Tuberc. Lung Dis. Off. J. Int. Union Tuberc. Lung Dis. 21, 965–976 (2017).

30. Jo, Y. et al. Model-based Cost-effectiveness of State-level Latent Tuberculosis Interventions in California, Florida, New York, and Texas. Clin. Infect. Dis. Off. Publ. Infect. Dis. Soc. Am. 73, e3476–e3482 (2021).

31. Tasillo, A. et al. Cost-effectiveness of Testing and Treatment for Latent Tuberculosis Infection in Residents Born Outside the United States With and Without Medical Comorbidities in a Simulation Model. JAMA Intern. Med. 177, 1755–1764 (2017).

32. Swartwood, N. A. et al. Tabby2: a user-friendly web tool for forecasting state-level TB outcomes in the United States. BMC Med. 21, 331 (2023).

33. Goodell, A. J. et al. Outlook for tuberculosis elimination in California: An individual-based stochastic model. PLoS ONE 14, e0214532 (2019).

34. Meaza, A. et al. Tuberculosis among refugees and migrant populations: Systematic review. PLoS ONE 17, e0268696 (2022).

35. Proença, R. et al. Active and latent tuberculosis in refugees and asylum seekers: a systematic review and meta-analysis. BMC Public Health 20, 838 (2020).

36. Pépin, J. et al. Impact and benefit-cost ratio of a program for the management of latent tuberculosis infection among refugees in a region of Canada. PloS One 17, e0267781 (2022).

37. Marx, F. M., Hauer, B., Menzies, N. A., Haas, W. & Perumal, N. Targeting screening and treatment for latent tuberculosis infection towards asylum seekers from high-incidence countries – a model-based cost-effectiveness analysis. BMC Public Health 21, 2172 (2021).

38. Wingate, L. T. et al. A cost-benefit analysis of a proposed overseas refugee latent tuberculosis infection screening and treatment program. BMC Public Health 15, 1201 (2015).

39. Campbell, J. R. et al. Cost-effectiveness of Latent Tuberculosis Infection Screening before Immigration to Low-Incidence Countries. Emerg. Infect. Dis. 25, 661–671 (2019).

40. Dale, K. D., Abayawardana, M. J., McBryde, E. S., Trauer, J. M. & Carvalho, N. Modeling the Cost-Effectiveness of Latent Tuberculosis Screening and Treatment Strategies in Recent Migrants to a Low-Incidence Setting. Am. J. Epidemiol. 191, 255–270 (2022).

41. Coker, R. & van Weezenbeek, K. L. Mandatory screening and treatment of immigrants for latent tuberculosis in the USA: just restraint? Lancet Infect. Dis. 1, 270–276 (2001).

42. Institute of Medicine (US) Committee on the Elimination of Tuberculosis in the United States. Ending Neglect: The Elimination of Tuberculosis in the United States. (National Academies Press (US), Washington (DC), 2000).

43. Berrocal-Almanza, L. C. et al. Effectiveness of nationwide programmatic testing and treatment for latent tuberculosis infection in migrants in England: a retrospective, population-based cohort study. Lancet Public Health 7, e305–e315 (2022).

44. Beeler Asay, G. R., et al. Cost-effectiveness of expanded latent TB infection testing and treatment: Lynn City, Massachusetts, USA. Int. J. Tuberc. Lung Dis. Off. J. Int. Union Tuberc. Lung Dis. 28, 21–28 (2024).

45. Yuen, C. M., Kammerer, J. S., Marks, K., Navin, T. R. & France, A. M. Recent Transmission of Tuberculosis - United States, 2011-2014. PloS One 11, e0153728 (2016).

