## Supplementary Appendix for "Cost-effectiveness of overseas testing and treatment for tuberculosis infection among United States-bound refugees: a mathematical modelling analysis"

**Table S1. Countries included in the analysis**

| **Countries of Birth (ISO 3166 alpha-3[1])** |
|---|
| AFG, AGO, ARE, ARM, AZE, BDI, BEL, BEN, BFA, BGD, BGR, BIH, BLR, BRA, BRN, BTN, BWA, CAF, CHN, CIV, CMR, COD, COG, COL, CUB, CYP, CZE, DEU, DJI, DOM, DZA, ECU, EGY, ERI, EST, ETH, FJI, FRA, FSM, GAB, GBR, GEO, GHA, GIN, GMB, GTM, GUM, HND, HTI, IDN, IND, IRN, IRQ, ISL, ISR, ITA, JOR, KAZ, KEN, KGZ, KHM, KOR, KWT, LAO, LBN, LBR, LBY, LKA, LTU, LVA, MAR, MDA, MDG, MEX, MLI, MLT, MMR, MNG, MOZ, MRT, MWI, MYS, NAM, NER, NGA, NIC, NPL, NRU, PAK, PAN, PER, PHL, PNG, POL, PRK, QAT, ROU, RUS, RWA, SAU, SDN, SEN, SLE, SLV, SMR, SOM, SSD, STP, SWE, SYR, TCD, TGO, THA, TJK, TKM, TUN, TUR, TZA, UGA, UKR, URY, UZB, VEN, VNM, YEM, ZAF, ZMB, ZWE<br>(A total of 128 countries) |
| **Countries of Examination (ISO 3166 alpha-3[1])** |
| AFG, AGO, ARG, ARM, AZE, BDI, BEN, BFA, BGD, BIH, BLR, BRA, BRN, BWA, CAF, CHN, CIV, CMR, COD, COG, COL, DJI, DOM, DZA, ECU, ERI, ETH, FJI, GAB, GEO, GHA, GIN, GMB, GTM, GUY, HKG, HND, HTI, IDN, IND, IRQ, KAZ, KEN, KGZ, KHM, KOR, LAO, LBR, LKA, LTU, MAC, MAR, MDA, MDG, MEX, MLI, MMR, MNG, MOZ, MRT, MWI, MYS, NAM, NER, NGA, NIC, NPL, NRU, PAK, PER, PHL, PLW, PNG, QAT, ROU, RUS, RWA, SDN, SEN, SLE, SLV, SOM, SSD, TCD, TGO, THA, TJK, TKM, TLS, TUN, TZA, UGA, UKR, URY, UZB, VEN, VNM, YEM, ZAF, ZMB, ZWE<br>(A total of 101 countries) |

**Text S1. Countries (ISO 3166 alpha-3[1]) where Strategy 3 could not be implemented**

Based on our discussion with experts at the International Organization of Migration, we assumed that no LTBI treatment is able to be offered in the following countries in Strategy 3. That is, in Strategy 3, refugees from these countries shared the same simulated outcome as they would in Strategy 2, where LTBI testing was required prior to U.S. entry but no LTBI treatment was offered overseas: AGO, BEN, BWA, CIV, COD, COG, DJI, ERI, GAB, GMB, LBR, MDG, MLI, MOZ, MRT, NAM, NER, SDN, SEN, SOM, SSD, TCD, TCD, ZAF.

### Figure S1A: Flowchart for Strategy 1 (adults)*

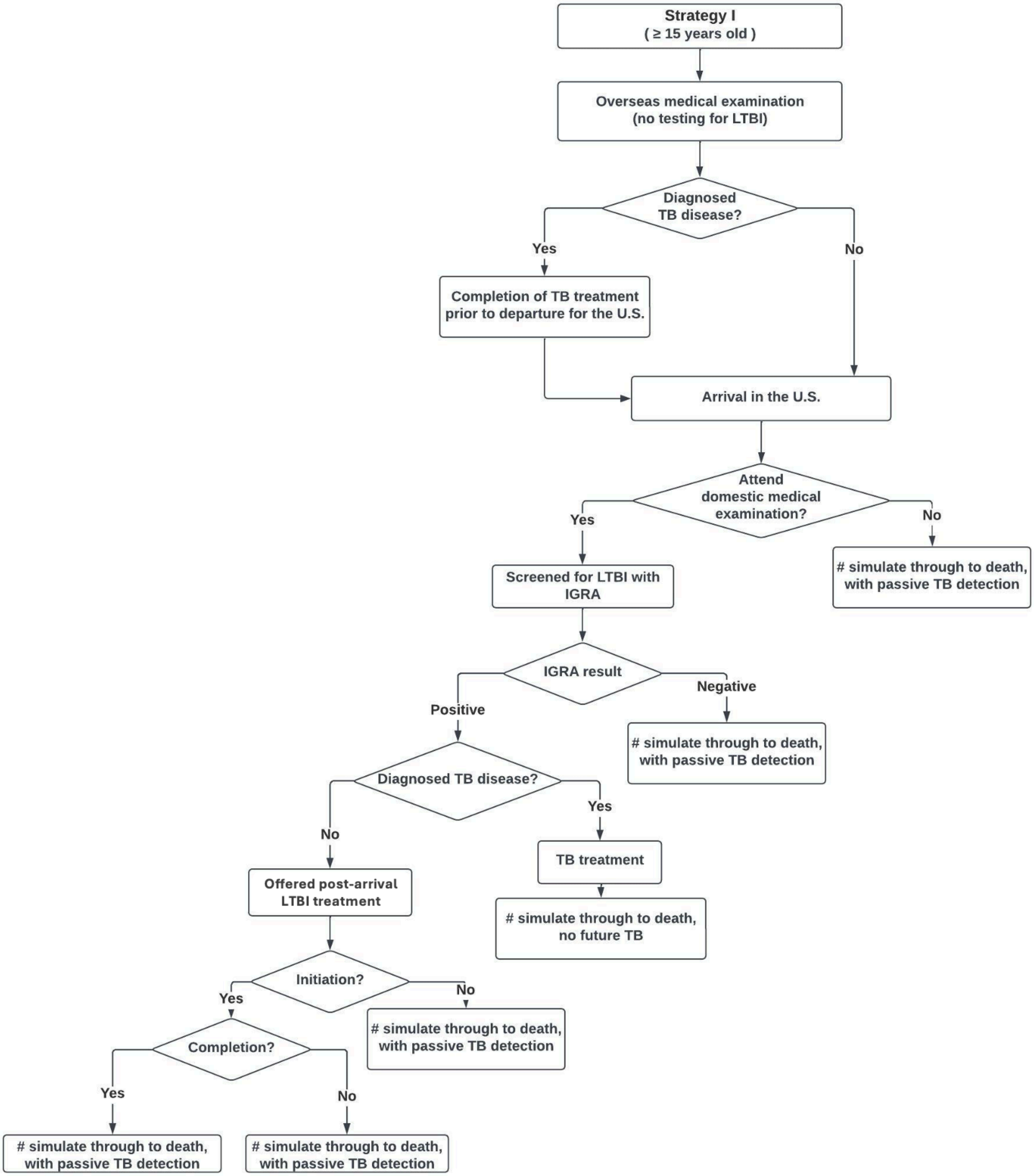


Abbreviations: LTBI, latent TB infection; U.S., United States; IGRA, interferon gamma release assay. * IGRA testing and LTBI treatment are offered after TB disease has been ruled out at the post-arrival medical examination. This strategy represents the pre-2024 screening requirements for U.S.-bound refugees ≥15 years old, tested in high-risk countries.

### Figure S1B: Flowchart for Strategy 2 (adults)*

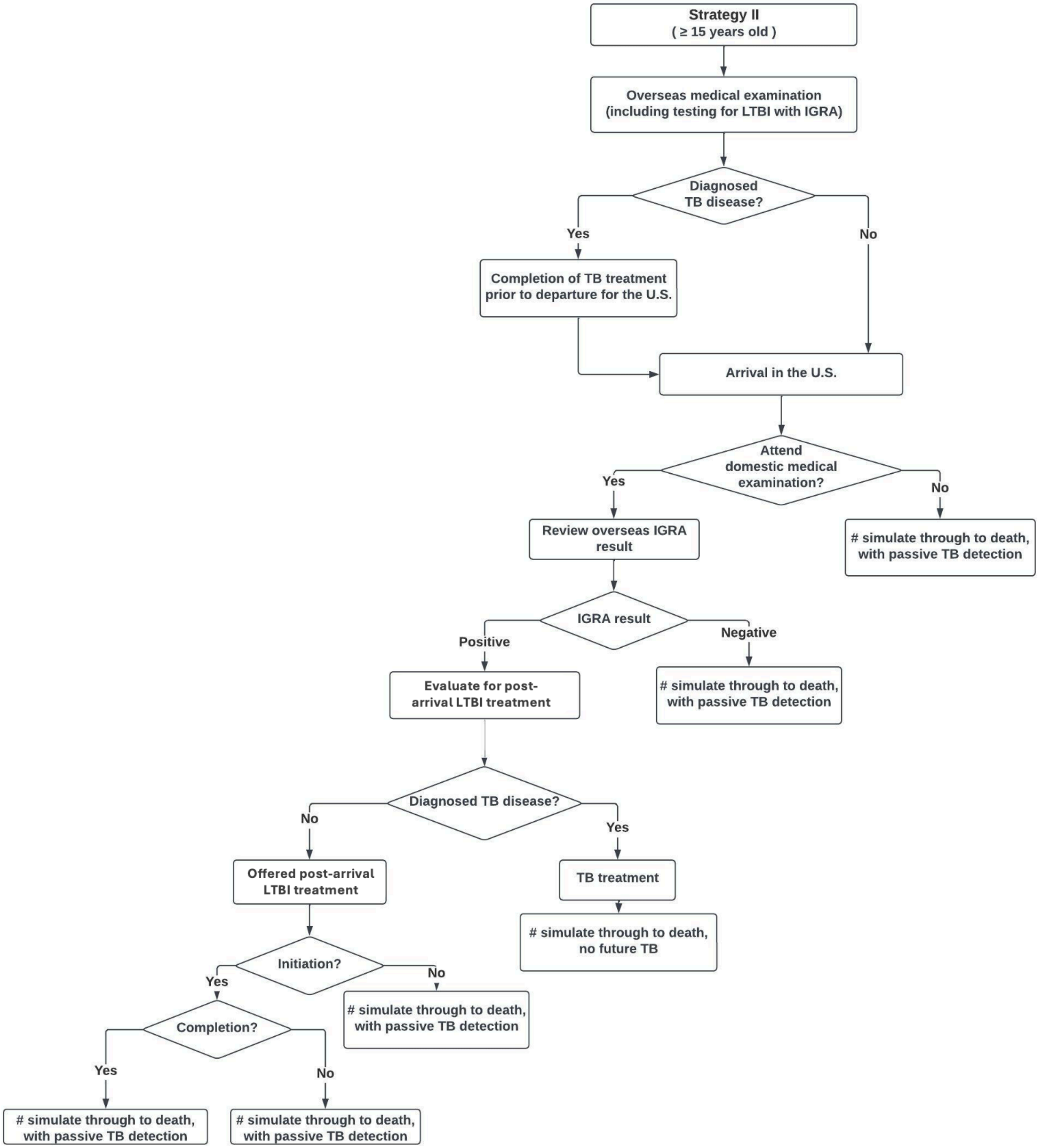


Abbreviations: LTBI, latent TB infection; U.S., United States; IGRA, interferon gamma release assay. * Pre-departure IGRA testing is required. Individuals testing IGRA-positive are offered LTBI treatment after TB disease has been ruled out at the post-arrival medical examination. This strategy represents the current screening requirements for U.S.-bound refugees ≥15 years old tested in high-risk countries, adopted in 2024.

### Figure S1C: Flowchart for Strategy 3 (adults)*

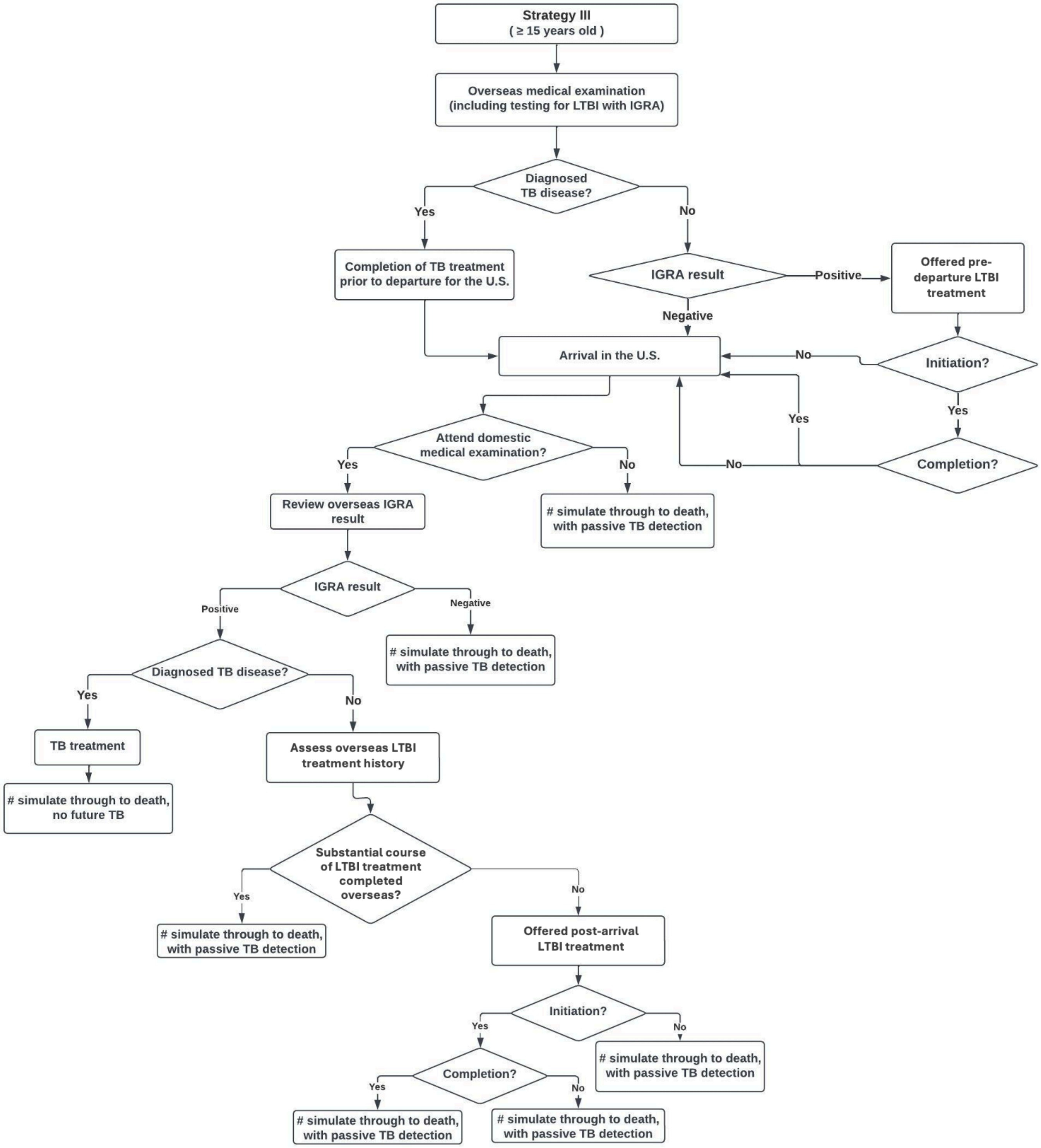


Abbreviations: LTBI, latent TB infection; U.S., United States; IGRA, interferon gamma release assay. * Pre-departure IGRA testing is required, with pre-departure LTBI treatment offered to individuals testing IGRA-positive, after TB disease has been ruled out. After U.S. arrival and domestic medical examination to rule out TB, LTBI treatment is offered again to those who decline LTBI treatment overseas. For those who initiate treatment overseas but do not complete the regimen, a proportion are assumed to re-initiate a full course of LTBI treatment during their post-arrival medical examination.

### Figure S1D: Flowchart for Strategies 1 and 2 (children)*

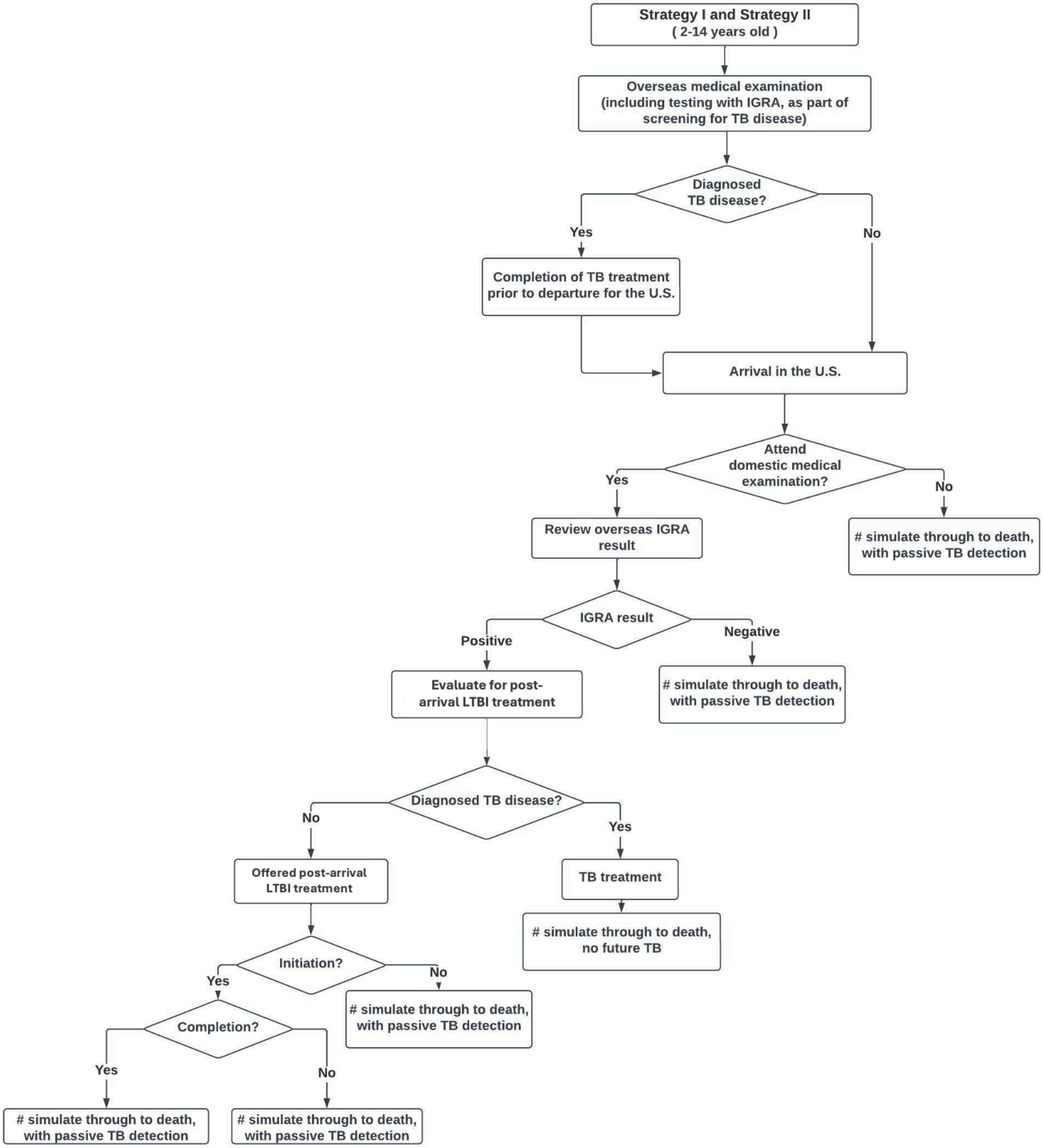


Abbreviations: LTBI, latent TB infection; U.S., United States; IGRA, interferon gamma release assay. * Pre-departure IGRA testing is required. Individuals testing IGRA-positive receive a medical examination after U.S. arrival to identify TB disease, with LTBI treatment offered after TB has been ruled out. This strategy represents the pre-2024 screening requirements for U.S.-bound refugees aged 2-14 years old tested in high-risk countries.

### Figure S1E: Flowchart for Strategy 3 (children)*

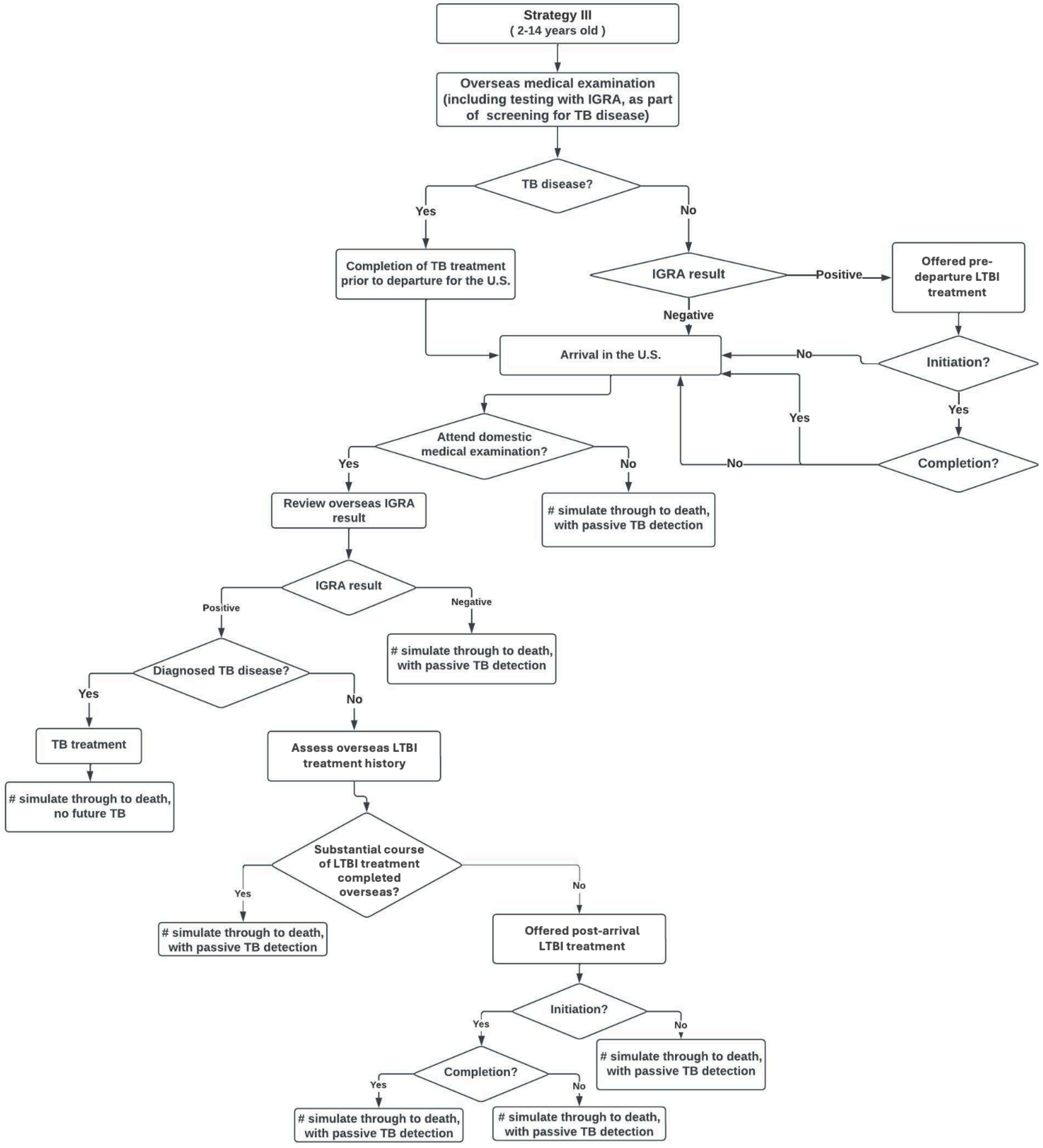


Abbreviations: LTBI, latent TB infection; U.S., United States; IGRA, interferon gamma release assay. * Pre-departure IGRA testing is required, with pre-departure LTBI treatment offered to individuals testing IGRA-positive, after TB disease has been ruled out. After U.S. arrival and domestic medical examination to rule out TB, LTBI treatment is offered again to those who decline LTBI treatment overseas. For those who initiate treatment overseas but do not complete the regimen, a proportion are assumed to re-initiate a full course of LTBI treatment during their post-arrival medical examination.

### Table S2. Parameter Table

| Parameter | Point estimate (95% interval, if included in probabilistic sensitivity analysis) | Source and comments |
|---|---|---|
| **Epidemiological parameters** | | |
| Background mortality rate | Varies by age | 2017 US Life Tables for foreign born individuals [2] |
| Mortality hazard ratio among those completed TB treatment | 1.78 | Following Lee Rodriguez 2020 *JAMA Netw Open* [3] , the hazard ratio was applied for 6 years after completion of TB treatment. |
| LTBI prevalence | Varies by age group and country-of-birth | Menzies 2024 *Lancet Global Health* [4] |
| Future TB incidence rates for the study cohort | Varies by age, country-of-birth, and time since entry | Hill 2020 *Epidemics* [5] |
| **Overseas IGRA testing and LTBI treatment (when implemented, see Figure S1)** | | |
| % Screened with IGRA | 100 | Assumption |
| % Initiated LTBI treatment,(3HP) conditional on a positive IGRA result | 65 (51, 79) | Assumption |
| % Completed 3HP regimen, or completed a sufficient proportion of regimen prior to U.S. entry such that re-initiation is not required, conditional on having initiated treatment | 88 (83, 92) | Assumption |
| **Domestic IGRA testing and LTBI treatment (when implemented, see Figure S1)** | | |
| % Attended domestic medical evaluation for refugees | 88 (83, 92) | Assumption |
| % Screened with IGRA, conditional on not tested with IGRA overseas and no signs of TB disease | 100 | Assumption |
| % Offered LTBI treatment (3HP) domestically, conditional on a positive IGRA result from overseas or domestic testing, with TB disease ruled out and one of the following: not offered LTBI treatment overseas, offered but did not initiate LTBI treatment overseas, or initiated overseas but did not complete a sufficient proportion of LTBI treatment signaling a need to re-initiate treatment | 100 | Assumption |
| % Initiated 3HP, conditional on being offered treatment, regardless of LTBI treatment history | 76 (69, 82) | CDC data from the Electronic Disease Notification 2013-2023, provided by Christina Phares, Division of Global Migration Health, 2024-03-20. (variables: DiagnosisBest, USTreatmentInitYesBest) [6] |

| Parameter | Point estimate (95% interval, if included in probabilistic sensitivity analysis) | Source and comments |
|---|---|---|
| % Completed 3HP regimen, conditional on having initiated treatment, regardless of LTBI treatment history | 63 (56, 69) | CDC data from the Electronic Disease Notification 2013-2023, provided by Christina Phares, Division of Global Migration Health, 2024-03-20. (variables: USTreatmentInitYesBest, USTreatmentComp) [6] |
| Time from U.S. entry to post-arrival domestic medical evaluation, if attended (months) | 1.0 | CDC data from the Electronic Disease Notification 2013-2023, provided by Christina Phares, Division of Global Migration Health, 2024-03-20. (variable: US_ArrivalToExam) [6] |
| **Test characteristics** | | |
| Specificity of IGRA for LTBI (%) | | |
| Aged < 15 years (based on proportion IGRA-negative among individuals with no TB disease) | 96 (84, 97) | Hirabayashi 2024 JIC [7] |
| Aged ≥ 15 years (based on proportion IGRA-negative among individuals with no evidence of TB exposure) | 98 (95, 99) | Jonas 2023 JAMA Network [8] |
| Sensitivity of IGRA for LTBI (%) | | |
| Aged < 15 years (based on proportion IGRA-positive among individuals with TB disease) | 77 (43, 94) | Hirabayashi 2024 JIC [7] |
| Aged ≥ 15 years (based on proportion IGRA-positive among individuals with TB disease) | 89 (84, 94) | Jonas 2023 JAMA Network [8] |
| Specificity of TB diagnosis (%) | 100 | Assumption |
| Sensitivity of TB diagnosis (%) | 100 | Assumption |
| **Treatment characteristics** | | |
| ***LTBI treatment*** | | |
| Length of LTBI treatment received in the U.S. among those who initiated a new course of LTBI treatment in the U.S. (regardless of LTBI treatment history overseas), and completed the regimen (weeks) | 18.2 | Feng 2023 J. Clin. Tuberc. Mycobact. Dis.[9] |
| Length of LTBI treatment received in the U.S. among those who initiated a new course of LTBI treatment in the U.S. (regardless of LTBI treatment history overseas), and did not complete the regimen (weeks) | 3.0 | Assumption |
| % Reduction in risk of future TB disease due to LTBI (efficacy), for individuals not completing LTBI treatment | 0 | Assumption |

| Parameter | Point estimate (95% interval, if included in probabilistic sensitivity analysis) | Source and comments |
|---|---|---|
| % Reduction in risk of future TB disease due to LTBI (efficacy), for individuals completing LTBI treatment | 93 | Sterling 2011 NEJM [10] |
| ***TB treatment (HRZE)*** | | |
| % Initiated treatment for TB disease, among those diagnosed with TB disease via active screening or through passive detection | 100 | Assumption |
| % Dead at TB diagnosis and therefore not eligible to begin TB treatment, among those diagnosed with TB disease | 1.6 | Unpublished CDC estimates from surveillance data for non-US–born population, National Tuberculosis Surveillance System, 2010-2019. [11] |
| % Completed treatment for TB, among those who do not die on TB treatment | 91.6 | |
| % Died on TB treatment, among those alive at TB diagnosis | 5.4 | |
| Length of TB treatment received in the U.S. among those who initiated TB treatment in the U.S. and completed the treatment (weeks) | 36.0 | Assumption based on 6-months treatment for drug-susceptible or drug-resistant TB |
| Length of TB treatment received in the U.S. among those who initiated TB treatment in the U.S. and did not complete the treatment (weeks) | 3.0 | Assumption |
| Time between symptom onset and diagnosis for TB cases not identified through the post-arrival medical examination (year) | 0.5 | Assumption |
| Time between TB diagnosis and treatment initiation (days) | 3.0 | Assumption |
| Time before treatment reinitiation, after treatment discontinuation (months) | 6.0 | Assumption |
| % Cured of TB, among individuals not completing TB treatment | 0 | Assumption |
| % Cured of TB, among individuals completing TB treatment | 100 | Assumption |
| **Utility weights** | | |
| No LTBI | 1.0 | |
| LTBI not on treatment | 1.0 | |
| On LTBI treatment | No toxicity: 1.0<br>With toxicity: 0.75 | Jo 2021 *Clin Infect Dis Off Publ Infect Dis Soc Am* [12] |
| TB disease, not receiving treatment | 0.75 (0.66, 0.83) | Bauer et al, *Qual Life Res* 2015 [13] |
| TB disease, on TB treatment | Untreated TB, 1st month of treatment: 0.75 (0.66, 0.83)<br>TB treatment after 1st month: 0.89 (0.76, 0.95) | Bauer et al, *Qual Life Res* 2015 [13] |

| Parameter | Point estimate (95% interval, if included in probabilistic sensitivity analysis) | Source and comments |
|---|---|---|
| Individuals surviving TB disease | 0.99 (0.981, 0.993) | Menzies 2021 *Lancet GH* [14] |
| **Cost parameters for overseas TB services (2023 US dollars)** | | |
| Cost of IGRA, incl. all costs (per test) | 50 (29, 77) | IOM estimates |
| Cost of LTBI treatment (3HP, DOT) for those who initiated treatment and received the full course of treatment | | GDF and IOM estimates, see Tables S4-1, S4-2 |
| Adults | 214 (105, 361) | |
| Children | 208 (99, 357) | |
| Cost of LTBI treatment (3HP, DOT) for those who initiated treatment and discontinued treatment prior to entering U.S. | | GDF and IOM estimates, assuming a discontinuation after a mean of 3 visits. |
| Adults | 53 (35, 74) | |
| Children | 52 (34, 74) | |
| **Cost parameters for domestic TB services (2023 US dollars)** | | |
| Cost of IGRA, incl. all costs (per test) | 61.98 (47, 79) | CMS Clinical Lab Fee Schedule 2023 [15] |
| Cost of TB screening with chest x-ray 2 views, incl. all costs (per test) | 34.23 (25, 45) | CMS Physician Fee Schedule 2023 [16] |
| Cost of LTBI treatment (3HP, SAT) for those who initiated treatment (same for cases of treatment completion and discontinuation) | | CDC estimates for LTBI treatment costs 2021, see Tables S4-1, S4-2 |
| Adults | 486 (275, 756) | |
| Children | 528 (314, 799) | |
| Cost of TB treatment for those who initiated treatment (same for cases of treatment completion and non-completion) | 24,846 (16,115, 35, 419) | Winston 2023 *EID* [17] |
| Annual discount rate (%) | 3.0 | Neumann 2016 Cost-Effectiveness in Health and Medicine [18] |
| Personal Consumption Expenditure Health (PCE-Health) Index | Varies by year | US Bureau of Economic Analysis [19] |

Abbreviations: LTBI, latent TB infection; U.S., United States; IGRA, interferon gamma release assay; DOT, directly observed therapy; SAT, self-administered therapy; HRZE, standard TB treatment regimen (isoniazid, rifampicin, pyrazinamide, ethambutol); 3HP, once-weekly isoniazid and rifapentine for 3 months.

### Table S3. Distributional characteristics of the uncertain input parameters for probabilistic sensitivity analysis

| Parameters | Mean | Median | 95% Uncertainty Interval | Parameter Distribution | *Source and comments* |
|---|---|---|---|---|---|
| **Overseas TBI testing and treatment** | | | | | |
| % Initiated LTBI treatment, conditional on a positive IGRA result | 65.0 | 65.0 | 50.8 – 79.3 | Uniform (0.50, 0.80) | IOM estimates |
| % Completed the full course of LTBI treatment, or completed a sufficient proportion of LTBI treatment prior to U.S. entry such that a re-initiation is not required, conditional on having initiated LTBI treatment | 88.0 | 88.2 | 82.7 – 92.5 | Beta (147.80, 20.16) | Khan 2022 *EID* [20] |
| **Domestic TBI testing and treatment** | | | | | |
| % Attended domestic medical evaluation for refugees | 88.0 | 88.2 | 82.7 – 92.5 | Beta (147.80, 20.16) | Assumption |
| % Initiated LTBI treatment, conditional on being offered treatment, regardless of LTBI treatment history | 76.0 | 76.1 | 69.2 – 82.2 | Beta (124.00, 39.16) | CDC data from the Electronic Disease Notification 2013-2023, provided by Christina Phares, Division of Global Migration Health, 2024-03-20. [6] |
| % Completed LTBI treatment, conditional on having initiated treatment, regardless of LTBI treatment history | 63.0 | 63.0 | 56.4 – 69.4 | Beta (131.54, 77.25) | CDC data from the Electronic Disease Notification 2013-2023, provided by Christina Phares, Division of Global Migration Health, 2024-03-20. [6] |
| **Test characteristics** | | | | | |
| Specificity (%) of IGRA for < 15 years old, based on proportion IGRA-negative among individuals with no TB disease | 96.0 | 96.8 | 87.6 – 99.7 | Beta (274.45, 3.05) | Hirabayashi 2024 J Infect and Chemotherapy [7] |
| Specificity (%) of IGRA for ≥ 15 years old, (based on proportion IGRA-negative among individuals with no evidence of TB exposure | 98.0 | 98.2 | 95.6 – 99.5 | Beta (191.10, 3.90) | Jonas 2023 JAMA Network [8] |
| Sensitivity (%) of IGRA for < 15 years old, based on proportion IGRA-positive among individuals with TB disease | 77.0 | 78.9 | 47.8 – 96.0 | Beta (7.62, 2.28) | Hirabayashi 2024 J Infect and Chemotherapy [7] |
| Sensitivity (%) of IGRA for ≥ 15 years old, based on proportion IGRA-positive among individuals with TB disease | 89.0 | 89.2 | 83.7 – 93.4 | Beta (138.52, 17.12) | Jonas 2023 JAMA Network [8] |

| Parameters | Mean | Median | 95% Uncertainty Interval | Parameter Distribution | *Source and comments* |
|---|---|---|---|---|---|
| **Treatment characteristics** | | | | | |
| **TB treatment (HRZE)** | | | | | |
| Time between symptom onset and diagnosis for TB cases not identified through the post-arrival screening program (year) | 0.50 | 0.50 | 0.25 – 0.75 | Normal (0.5, 0.125) | Assumption of 6 months, with 3-9 month 95% uncertainty interval. |
| % Completed treatment for TB, among those that do not die on TB treatment | 91.6 | 91.7 | 89.0 – 93.9 | Beta (450.16, 41.28) | Unpublished CDC estimates from surveillance data for non-US–born population, National Tuberculosis Surveillance System, 2010-2019 [11] |
| **Utility weights** | | | | | |
| TB not on TB or LTBI treatment | 0.750 | 0.752 | 0.660 – 0.826 | 1 – Gamma (34.60, 0.007225)* | Bauer 2015 Qual Life Res [13] |
| Anyone on TB treatment (1st month of treatment) | 0.750 | 0.752 | 0.660 – 0.826 | 1 – Gamma (34.60, 0.007225) | Bauer 2015 Qual Life Res [13] |
| Anyone on TB treatment (after 1st month of treatment) | 0.874 | 0.880 | 0.761 – 0.951 | 1 – Gamma (6.61, 0.01906) | Bauer 2015 Qual Life Res [13] |
| Anyone after TB treatment completion if cured | 0.988 | 0.988 | 0.981 – 0.993 | 1 – Gamma (16, 0.00075) | Menzies 2021 *Lancet GH*[14] |
| **Cost parameters for overseas TB services** | | | | | |
| LTBI treatment (3HP, DOT) for those who initiated treatment and received the full course of treatment (2023 US dollars) for adults – healthcare costs | 214 | 207 | 104 – 363 | Gamma (10.30, 20.77) | GDF and IOM estimates, see Tables S4-2 |
| LTBI treatment (3HP, DOT) for those who initiated treatment and received the full course of treatment (2023 US dollars) for children – healthcare costs | 208 | 201 | 99 – 358 | Gamma (9.73, 21.37) | GDF and IOM estimates, see Tables S4-1 |
| LTBI treatment (3HP, DOT) for those who initiated treatment and discontinued treatment prior to entering U.S. (2023 US dollars) for adults – healthcare costs | 53 | 52 | 35 – 74 | Gamma (28.09, 1.89) | GDF and IOM estimates, assuming a discontinuation after a mean of 3 visits. |
| LTBI treatment (3HP, DOT) for those who initiated treatment and discontinued treatment prior to entering U.S. (2023 US dollars) for children – healthcare costs | 52 | 51 | 34 – 73 | Gamma (27.04, 1.92) | GDF and IOM estimates, assuming a discontinuation after a mean of 3 visits. |
| IGRA, incl. all costs (per test, 2023 US dollars) | 50 | 49 | 29 – 77 | Gamma (16.00, 3.13) | IOM estimates |
| **Cost parameters for domestic TB services** | | | | | |

| Parameters | Mean | Median | 95% Uncertainty Interval | Parameter Distribution | *Source and comments* |
|---|---|---|---|---|---|
| LTBI treatment (3HP, SAT) for those who initiated treatment (same for cases of treatment completion and discontinuation; 2023 US dollars) for adults – healthcare costs | 486 | 475 | 275 – 760 | Gamma (15.12, 32.15) | CDC LTBI treatment cost estimates, see Table S4-2 |
| LTBI treatment (3HP, SAT) for those who initiated treatment (same for cases of treatment completion and discontinuation; 2023 US dollars) for children – healthcare costs | 528 | 518 | 312 – 800 | Gamma (17.84, 29.59) | CDC LTBI treatment cost estimates, see Table S4-1 |
| TB disease treatment for those who initiated treatment (same for cases of treatment completion and non-completion) (2023 US dollars) – healthcare costs | 24,846 | 24,511 | 16,031 – 35,561 | Gamma (24.69, 1006.20) | Winston 2023 *EID* [17] |
| IGRA, incl. all costs (per test, 2023 US dollars) | 61.98 | 61.61 | 46.73 – 79.35 | Gamma (55.32, 1.12) | CMS Clinical Lab Fee Schedule 2023 [15] |
| TB screening with chest x-ray 2 views, incl. all costs (per test, 2023 US dollars) | 34.23 | 33.99 | 25.14 – 44.70 | Gamma (46.87, 0.73) | CMS Physician Fee Schedule 2023 [16] |

Abbreviations: LTBI, latent TB infection; U.S., United States; IGRA, interferon gamma release assay; DOT, directly observed therapy; SAT, self-administered therapy; HRZE, standard TB treatment regimen (isoniazid, rifampicin, pyrazinamide, ethambutol); 3HP, once-weekly isoniazid and rifapentine for 3 months. * Gamma distribution parameters are expressed as shape and scale.

**Table S4-1.** Cost estimation for complete course of LTBI treatment overseas and in the United States, for children

| Categories | Overseas | Domestic |
|---|---|---|
| **LTBI treatment regimen** | 12 weeks of once-weekly isoniazid (900 mg) and rifapentine (900 mg), directly observed therapy [3HP, DOT] | 12 weeks of once-weekly isoniazid (900 mg) and rifapentine (600 mg), self-administered therapy [3HP, SAT] |
| Total cost for medication | $11 | $171 |
| Weekly cost for medication | $0.94 | $14.28 |
| Total doses | 12 | 12 |
| Total cost for clinic visits | $180 | $225 |
| Total clinic visits | 12 | 3 |
| Unit cost for the initial clinic visit | $15 | $156 |
| Unit cost for a follow-up clinic visit | $15 | $34 |
| Total supplies costs | $11 | $87 |
| Adverse events costs | $6 | $21 |
| ***Estimated total cost*** | ***$208*** | ***$528*** |

Abbreviations: LTBI, latent TB infection. Dosage for isoniazid and rifapentine in children is based on weight.[21] We used the mean weight of the sampled study population ≤ 14 (31.45 kilograms) to approximate the cost of 3HP. The mean weight was based on the mean age of the sampled children cohort (8 years old).[22] Estimates of domestic costs are based on by Veteran Affairs drug cost information, adjusted to 2023 US dollars.[23] Overseas clinic and supply costs are based on expert input from the International Organization of Migration, and overseas medication costs are based on Global Drug Facility catalogue.[24]

**Table S4-2.** Cost estimation for complete course of LTBI treatment overseas and in the United States, for adults

| Categories | Overseas | Domestic |
|---|---|---|
| **LTBI treatment regimen** | **12 weeks of once-weekly isoniazid (900 mg) and rifapentine (900 mg), directly observed therapy [3HP, DOT]** | **12 weeks of once-weekly isoniazid (900 mg) and rifapentine (900 mg), self-administered therapy [3HP, SAT]** |
| Total cost for medication | $17 | $130 |
| Weekly cost for medication | $1.38 | $10.85 |
| Total doses | 12 | 12 |
| Total cost for clinic visits | $180 | $224 |
| Total clinic visits | 12 | 3 |
| Unit cost for the initial clinic visit | $15 | $156 |
| Unit cost for a follow-up clinic visit | $15 | $34 |
| Total supplies costs | $12 | $87 |
| Adverse events costs | $6 | $43 |
| ***Estimated total cost*** | ***$214*** | ***$486*** |

Abbreviations: LTBI, latent TB infection. Estimates of domestic costs are based on by Veteran Affairs drug cost information, adjusted to 2023 US dollars.[23] Overseas clinic and supply costs are based on expert input from the International Organization of Migration, and overseas medication costs are based on Global Drug Facility catalogue.[24]

**Table S5-1.** Cost-effectiveness analysis results for the scenario analysis where LTBI prevalence is ***double the value assumed in the main analysis***

| Strategy | Total QALY per refugee (95% UI) | Total cost per refugee (95% UI) | Incremental QALYs (95% UI) | Incremental Costs (95% UI) | ICER ($ / QALY gained) |
|---|---|---|---|---|---|
| **Children** | | | | | |
| *No intervention scenario* | 29.8808 (29.8802 – 29.8813) | 51 (35 – 70) | -- | -- | -- |
| *Strategy 1, 2* | 29.8813 (29.8808 – 29.8817) | 106 (59 – 169) | -- | -- | Dominated by Strategy 3 |
| *Strategy 3* | 29.8816 (29.8811 – 29.8819) | 74 (33 – 136) | 0.0008 (0.0008 – 0.0013) | 23 (-2.2 – 54.7) | 28,750 |
| **Adults** | | | | | |
| *No intervention scenario* | 25.5133 (25.5115 – 25.5148) | 216 (161 – 282) | -- | -- | -- |
| *Strategy 1* | 25.5165 (25.5152 – 25.5177) | 365 (229 – 531) | -- | -- | Dominated by Strategy 3 |
| *Strategy 2* | 25.5165 (25.5152 – 25.5176) | 378 (235 – 564) | -- | -- | Dominated by Strategy 3 |
| *Strategy 3* | 25.5187 (25.5175 – 25.5196) | 275 (169 – 416) | 0.0054 (0.0042 – 0.0068) | 59 (-25.7 – 140) | 10,925 |

Abbreviations: LTBI, latent TB infection; QALY, quality-adjusted life year; 3HP once-weekly isoniazid and rifapentine for 3 months; UI, uncertainty interval; --, not applicable. All costs and QALYs are discounted and in 2023 US dollars, analyzed from TB services perspective (overseas TB screening not included) with a lifetime analytic horizon. Costs are rounded to integers.
No intervention scenario: No IGRA testing and LTBI treatment for the refugee cohort, except for IGRA testing as part of mandatory TB screening in children. Strategy 1: pre-departure IGRA testing for children (2-14 years) and post-arrival IGRA testing for adults (>14 years), with 3HP offered in the United States for IGRA-positive individuals after ruling out TB disease Strategy 2: pre-departure IGRA testing children and adults, and post-arrival 3HP offered for IGRA-positive individuals after ruling out TB disease; Strategy 3: pre-departure IGRA testing for children and adults, and pre-departure 3HP offered for IGRA-positive individuals testing after ruling out TB disease, with 3HP re-offered in the United States for individuals not completing treatment before U.S. arrival.

**Table S5-2.** Cost-effectiveness analysis results for the scenario analysis where LTBI prevalence is ***half the value assumed in the main analysis***

| Strategy | Total QALY (per refugee, 95% UI) | Total cost (per refugee, 95% UI) | Incremental QAL` | Incremental cost | ICER ($ / QALY gained) |
|---|---|---|---|---|---|
| **Children** | | | | | |
| *No intervention scenario* | 29.8819 (29.8817 – 29.8821) | 12 (6 – 20) | -- | -- | -- |
| *Strategy 1, 2* | 29.8820 (29.8818 – 29.8822) | 36 (15– 77) | -- | -- | Dominated by Strategy 3 |
| *Strategy 3* | 29.8821 (29.8819 – 29.8822) | 28 (9 – 66) | 0.0002 (0.0002 – 0.0004) | 16 (0.3 – 46) | 80,000 |
| **Adults** | | | | | |
| *No intervention scenario* | 25.5201 (25.5194 – 25.5206) | 54 (38 – 73) | -- | -- | -- |
| *Strategy 1* | 25.5209 (25.5204 – 25.5213) | 122 (56 – 198) | -- | -- | Dominated by Strategy 3 |
| *Strategy 2* | 25.5209 (25.5204 – 25.5213) | 135 (80 – 208) | -- | -- | Dominated by Strategy 3 |
| *Strategy 3* | 25.5214 (25.5210 – 25.5217) | 109 (66 – 165) | 0.0013 (0.0009 – 0.0018) | 55 (23 – 89) | 42,308 |

Abbreviations: LTBI, latent TB infection; QALY, quality-adjusted life year; 3HP once-weekly isoniazid and rifapentine for 3 months; UI, uncertainty interval; --, not applicable. All costs and QALYs are discounted and in 2023 US dollars, analyzed from TB services perspective (overseas TB screening not included) with a lifetime analytic horizon. Costs are rounded to integers.

No intervention scenario: No IGRA testing and LTBI treatment for the refugee cohort, except for IGRA testing as part of mandatory TB screening in children. Strategy 1: pre-departure IGRA testing for children (2-14 years) and post-arrival IGRA testing for adults (>14 years), with 3HP offered in the United States for IGRA-positive individuals after ruling out TB disease Strategy 2: pre-departure IGRA testing children and adults, and post-arrival 3HP offered for IGRA-positive individuals after ruling out TB disease; Strategy 3: pre-departure IGRA testing for children and adults, and pre-departure 3HP offered for IGRA-positive individuals testing after ruling out TB disease, with 3HP re-offered in the United States for individuals not completing treatment before U.S. arrival.

### Reference


1 ISO - ISO 3166 — Country Codes. ISO. https://www.iso.org/iso-3166-country-codes.html (accessed Feb 12, 2025).

2 Medina M, Sabo S, and Vespa J. Living Longer: Historical and Projected Life Expectancy in the United States, 1960 to 2060. U.S. Census Bureau, 2020 https://www.census.gov/content/dam/Census/library/publications/2020/demo/p25-1145.pdf.

3 Lee-Rodriguez C, Wada PY, Hung Y-Y, Skarbinski J. Association of Mortality and Years of Potential Life Lost With Active Tuberculosis in the United States. *JAMA Network Open* 2020; **3**: e2014481.

4 Menzies NA, Swartwood NA, Cohen T, *et al.* The long-term effects of domestic and international tuberculosis service improvements on tuberculosis trends within the USA: a mathematical modelling study. *Lancet Public Health* 2024; **9**: e573–82.

5 Hill AN, Cohen T, Salomon JA, Menzies NA. High-resolution estimates of tuberculosis incidence among non-U.S.-born persons residing in the United States, 2000–2016. *Epidemics* 2020; **33**: 100419.

6 Unpublished CDC data from the Electronic Disease Notification 2013-2023 (only data children 2-14 years, provided by Christina Phares, Division of Global Migration Health, 2024-03-20.

7 Hirabayashi R, Nakayama H, Yahaba M, Yamanashi H, Kawasaki T. Utility of interferon-gamma releasing assay for the diagnosis of active tuberculosis in children: A systematic review and meta-analysis. *Journal of Infection and Chemotherapy* 2024; **30**: 516–25.

8 Jonas DE, Riley SR, Lee LC, *et al.* Screening for Latent Tuberculosis Infection in Adults: Updated Evidence Report and Systematic Review for the US Preventive Services Task Force. *JAMA* 2023; **329**: 1495–509.

9 Feng P-JI, Horne DJ, Wortham JM, Katz DJ. Trends in tuberculosis clinicians' adoption of short-course regimens for latent tuberculosis infection. *J Clin Tuberc Other Mycobact Dis* 2023; **33**: 100382.

10 Sterling TR, Villarino ME, Borisov AS, *et al.* Three Months of Rifapentine and Isoniazid for Latent Tuberculosis Infection. *N Engl J Med* 2011; **365**: 2155–66.

11 Unpublished 2022 CDC data for non-US born population residing in the United States for less than 10 years (2010-2019), the U.S. CDC Surveillance Team, Division of Tuberculosis Elimination, 2022-10-06.

12 Jo Y, Shrestha S, Gomes I, *et al.* Model-based Cost-effectiveness of State-level Latent Tuberculosis Interventions in California, Florida, New York, and Texas. *Clin Infect Dis* 2021; **73**: e3476–82.

13 Bauer M, Ahmed S, Benedetti A, *et al.* The impact of tuberculosis on health utility: a longitudinal cohort study. *Qual Life Res* 2015; **24**: 1337–49.

14 Menzies NA, Quaife M, Allwood BW, *et al.* Lifetime burden of disease due to incident tuberculosis: a global reappraisal including post-tuberculosis sequelae. *The Lancet Global Health* 2021; **9**: e1679–87.

15 Clinical Laboratory Fee Schedule | CMS. https://www.cms.gov/medicare/medicare-fee-for-service-payment/clinicallabfeesched (accessed Feb 19, 2023).

16 Physician Fee Schedule | CMS. https://www.cms.gov/medicare/payment/fee-schedules/physician (accessed June 30, 2025).

17 Winston CA, Marks SM, Carr W. Estimated Costs of 4-Month Pulmonary Tuberculosis Treatment Regimen, United States. *Emerg Infect Dis* 2023; **29**: 2102–4.

18 Neumann PJ, Ganiats TG, Russell LB, Sanders GD, Siegel JE, editors. Cost-Effectiveness in Health and Medicine. Oxford University Press, 2016 DOI:10.1093/acprof:oso/9780190492939.001.0001.

19 BEA Interactive Data Application. https://apps.bea.gov/iTable/?reqid=19&step=3&isuri=1&select_all_years=0&nipa_table_list=2014&series=m&first_year=2020&last_year=2022&scale=-99&categories=underlying&thetable=&_gl=1*3rlonf*_ga*MTgwMjc2NTUwMC4xNzM2NDU3MTQy*_ga_J4698JNNFT*MTczNjQ1NzE0Mi4xLjEuMTczNjQ1NzYxMy41LjAuMA..#eyJhcHBpZCI6MTksInN0ZXBzIjpbMSwyLDMsM10sImRhdGEiOltbImNhdGVnb3JpZXMiLCJVbmRlcmx5aW5nIl0sWyJOSVBBX1RhYmxlX0xpc3QiLCIyMDE3Il0sWyJGaXJzdF9ZZWFyIiwiMjAyMCJdLFsiTGFzdF9ZZWFyIiwiMjAyMyJdLFsiU2NhbGUiLCItNiJdLFsiU2VyaWVzIiwiTSJdXX0= (accessed Jan 9, 2025).

20 Khan A, Phares CR, Phuong HL, *et al.* Overseas Treatment of Latent Tuberculosis Infection in US–Bound Immigrants. *Emerg Infect Dis* 2022; **28**: 582–90.

21 Sterling TR. Guidelines for the Treatment of Latent Tuberculosis Infection: Recommendations from the National Tuberculosis Controllers Association and CDC, 2020. *MMWR Recomm Rep* 2020; **69**. DOI:10.15585/mmwr.rr6901a1.

22 Vital and Health Statistics, Series 3, Number 46.

23 Logistics O of P Acquisition and. VA.gov | Veterans Affairs. https://www.va.gov/opal/nac/fss/pharmprices.asp (accessed June 10, 2025).

24 GDF Product Catalog | Stop TB Partnership. https://www.stoptb.org/what-we-do/facilitate-access-tb-drugs-diagnostics/global-drug-facility-gdf/products-catalog (accessed June 30, 2025).
